# Dominant negative *ADA2* mutations cause ADA2 deficiency in heterozygous carriers

**DOI:** 10.1101/2024.12.09.24317629

**Authors:** Marjon Wouters, Lisa Ehlers, Wout Van Eynde, Meltem Ece Kars, Selket Delafontaine, Verena Kienapfel, Mariia Dzhus, Rik Schrijvers, Petra De Haes, Sofie Struyf, Giorgia Bucciol, Yuval Itan, Alexandre Bolze, Arnout Voet, Anneleen Hombrouck, Leen Moens, Benson Ogunjimi, Isabelle Meyts

## Abstract

Human ADA2 deficiency (DADA2) is an inborn error of immunity with a broad clinical phenotype which encompasses vasculopathy including livedo racemosa and lacunar strokes, as well as hemato-immunological features. Diagnosis is based on the combination of decreased serum ADA2 activity and the identification of biallelic deleterious alleles in the *ADA2* gene. DADA2 carriers harbor a single pathogenic variant in *ADA2* and are mostly considered healthy and asymptomatic. However, some DADA2 carriers present a phenotype compatible with DADA2. Here, we report ten patients from seven kindreds presenting with a phenotype indicative of DADA2, in whom only a single pathogenic variant (p.G47R, p.G47V, p.R169Q, p.H424N) was identified. To test whether being heterozygote for specific variants could explain the patients’ phenotype, we investigated the effect of the ADA2 missense variants p.G47A, p.G47R, p.G47V, p.G47W, p.R169Q, p.E328K, p.T360A, p.N370K, p.H424N and p.Y453C on ADA2 protein expression, secretion and enzymatic activity. Functional studies indicate that they exert a dominant negative effect on ADA2 enzymatic activity, dimerization and/or secretion. At the molecular level, heterozygosity for these variants mimics what is observed in DADA2. We conclude that humans with heterozygous dominant negative missense variants in ADA2 are at risk of DADA2.

**Graphical abstract:** 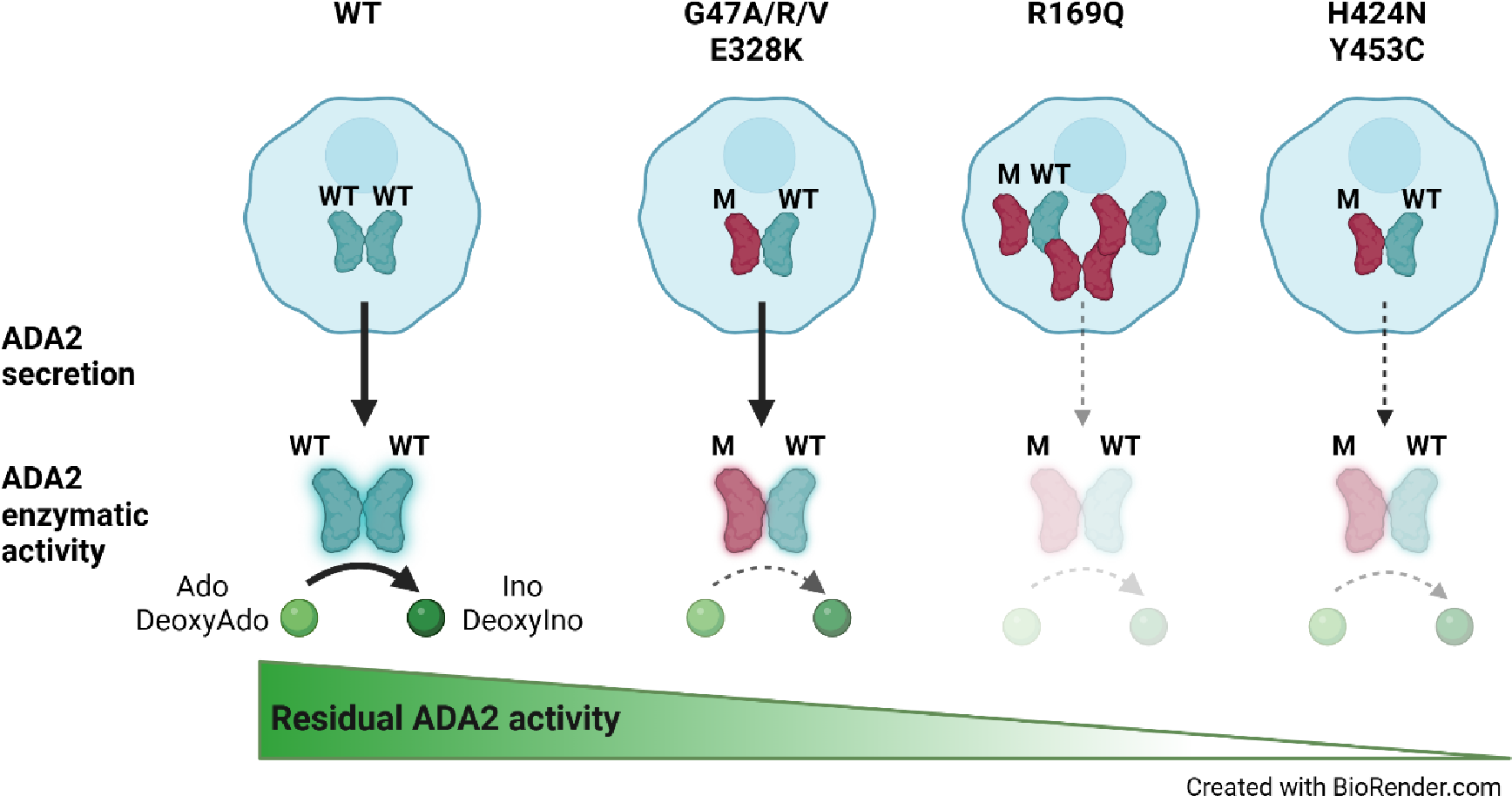

## Introduction

Human ADA2 deficiency (DADA2) is an inborn error of immunity (IEI) caused by biallelic deleterious mutations in *ADA2,* characterized by autoinflammation in the form of recurrent fevers and vasculitis ranging from livedo racemosa to lacunar strokes (1, 2). As additional patients have been described, the phenotype has expanded to include pure red cell aplasia, various forms of cytopenias and bone marrow failure. Furthermore, lymphoproliferation, hepatosplenomegaly, immunodeficiency with hypogammaglobulinemia, sinopulmonary and severe viral infections have been added to the phenotype (3, 4). DADA2 patients can also present with hematological malignancy and hemophagocytosis (5). To date, over 150 pathogenic variants in ADA2 have been described (6). *ADA2* encodes a 59 kD glycoprotein with a signal peptide and a dimerization domain. The theories on disease pathogenesis focus primarily on the reduction or absence of extracellular adenosine deaminase activity due to loss of ADA2 leading to a skewed macrophage development with predominance of proinflammatory M1 macrophages (7). The cytokine profile of patients is complex, featuring upregulation of both type I as well as type II interferons (IFNs) alongside to other proinflammatory cytokines (8–10). More recently, Greiner-Tollersrud et al. proposed that ADA2 functions as an adenosine deaminase acting on DNA in the lysosome, where it regulates immune sensing by modulating TLR9 activation (11). Regarding treatment, anti-TNF agents alleviate fever and vasculitis and can prevent strokes in most patients (12). Patients with hematological and immunological manifestations often require hematopoietic stem cell transplantation (13).

The diagnosis of DADA2 is based on decreased ADA2 serum enzyme activity and the identification of two deleterious alleles in the *ADA2* gene (14, 15). A model of genotype / phenotype correlation was proposed, identifying 25% residual adenosine deaminase activity as the pathogenicity threshold when the variant allele was tested in a HEK293T overexpression system (15, 16). In this model, the lowest residual ADA2 activity in the supernatant is associated with the most severe manifestations of DADA2, including pure red cell aplasia. The variants were tested in a homozygous state (15, 16). This model suggests that pathogenic variants which result in little or no residual activity, could be pathogenic in heterozygous state, especially since ADA2 functions as a dimer. Interestingly, extended immunophenotyping of carriers of DADA2 (i.e. harboring a single deleterious ADA2 allele) showed an intermediate phenotype for several of the features identified in DADA2 patients including SIGLEC-1 expression (8).

Diseased carriers of heterozygous variants have already been described in literature, however without mechanistic validation. In 2022, Moi. et al. reported a 35-year-old woman presenting with common variable immunodeficiency. Childhood medical history revealed large joint arthritis episodes and recurrent infection. Next-generation sequencing identified her as a heterozygous carrier for the R169Q variant in ADA2. She is the mother of a DADA2 patient, homozygous for the R169Q variant (17). Next, a study performed by the French reference center for autoinflammatory diseases investigated the DNA of 66 patients with clinically suspected DADA2. Three patients were found to be carrier of a heterozygous ADA2 variant. In particular, a heterozygous carrier of G47R presented with an inflammatory syndrome characterized by fever and increased CRP levels. Moreover, a papular rash with pruritis and gastrointestinal manifestations were observed (18). In addition, in a Japanese cohort study by Nihira et al. two siblings were reported to suffer from livedo racemosa, renal infarction and neurological manifestations (cerebral infarction and memory disturbance). Next-generation sequencing revealed the presence of a single pathogenic variant E328K (10). Finally, in 2023, Izumo et al. described a 13-year-old girl that was hospitalized with sudden-onset weakness in the right upper and lower limbs caused by a cerebral infarction. Four years earlier, she had presented with livedo. Her treatment included oral anti-platelet drugs and steroid pulse therapy. Additionally, splenic and renal infarction were noted. Measurement of serum ADA2 enzymatic activity revealed levels at 50% of healthy controls. She was diagnosed with polyarteritis nodosa and was treated with prednisone and cyclophosphamide (19). However, she developed a new cerebral infarction and was then started on infliximab. Since then, no reoccurrence of cerebral infarction has been observed. Family history revealed that her father had suffered from an acute subarachnoid hemorrhage in his 40s. Sanger sequencing identified the missense variants F355L and Y453C, without functional validation (19).

Our interest in the effect of heterozygous variants was further driven by the finding that in a HEK293T overexpression system, the heterozygous ADA2 condition, expressing 50% R169Q ADA2 and 50% wild-type (WT) ADA2 resulted in reduced total ADA2 protein secretion compared to the homozygous WT ADA2 condition (20). In addition, in clinical practice, we encountered several patients with a phenotype suggestive of DADA2, in whom we identified only a single deleterious allele.

In this report, we describe how specific heterozygous variants cause ADA2 deficiency according to the proposed pathogenicity cut-off (16) through distinct dominant negative effects on either ADA2 enzyme activity, dimerization and/or secretion.

## Results

### Clinical phenotype and genotype of reported and suspected DADA2 patients in whom only a heterozygous variant in ADA2 was identified

We identified ten patients, from seven kindreds, with a phenotype suggestive of DADA2, each harboring a single mutation in *ADA2,* in addition to the patients with heterozygous ADA2 variants retrieved from the literature (10, 17–19). Their pedigrees, clinical features and genetic characteristics are summarized in Figure. 1A, Supplemental Tables. 4-6 and Supplemental Figure. 1. All patients were born to non-consanguineous parents of either Moroccan or Belgian descent. Patient 1 (P1) is the son of patient 2 (P2) (kindred A), and patient 3 (P3) is the daughter of patient 4 (P4) (kindred B). For patient 5 (P5) (kindred C) and patient 6 (P6) (kindred D) parental DNA was unavailable. Patient 7 (P7) and 8 (P8) are the parents of two previously reported DADA2 patients (kindred E), and patient 9 (P9) is the father of two other previously reported DADA2 patients (kindred F). Parental DNA was also unavailable for patient 10 (P10).

**Figure 1.**
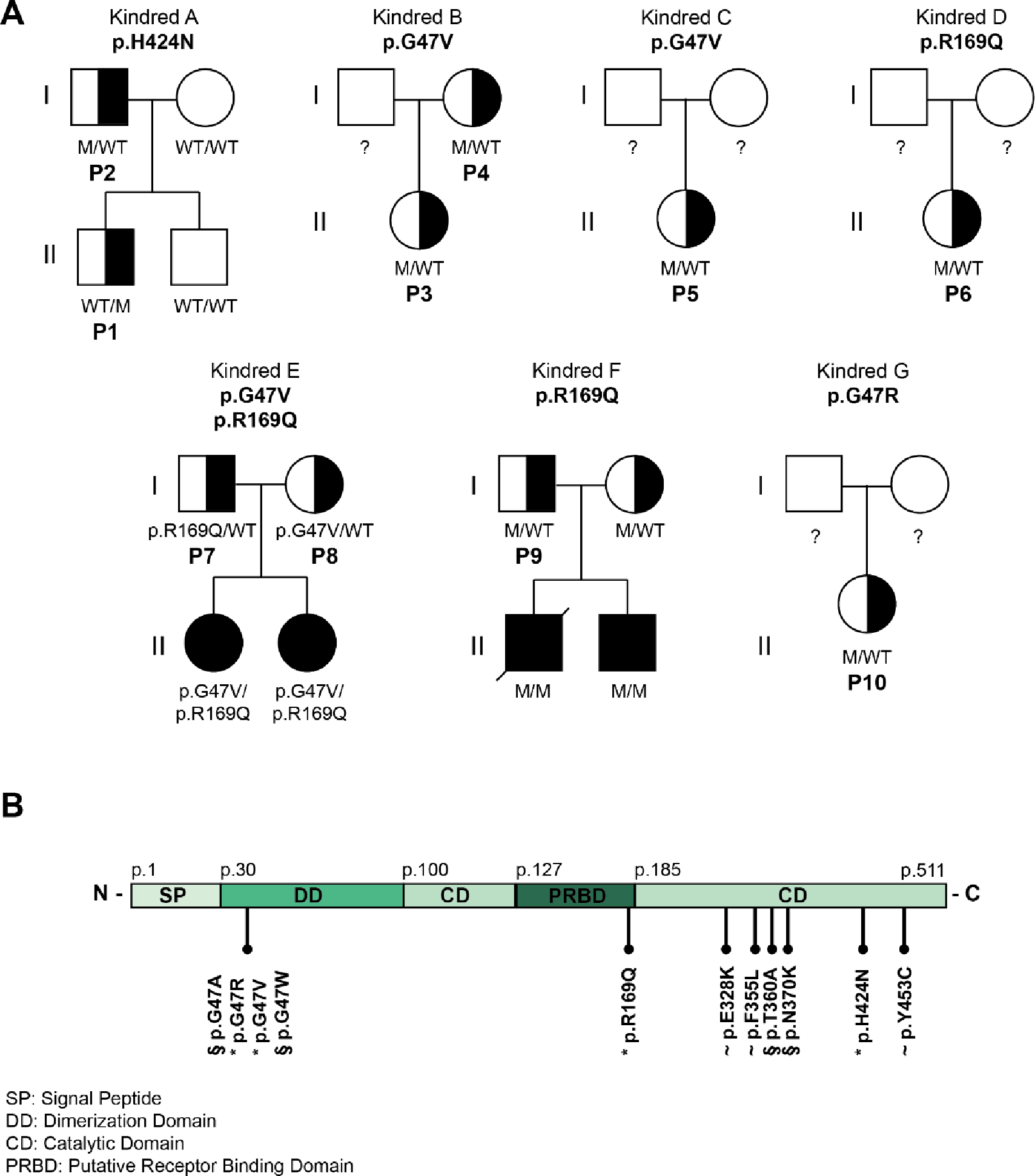
Pedigree analysis of 10 DADA2 carriers presenting with DADA2 clinical phenotype. **A.** Pedigrees of seven kindreds showing familial segregation of ADA2 missense variants. M represents mutant. Individuals with unknown genotype are labeled ‘?’. Black filled symbols represent individuals with 2 pathogenic alleles, half-filled symbols represent individuals with 1 pathogenic allele ‘P’ represents individuals carrying 1 pathogenic allele with a DADA2 phenotype. **B.** Schematic representation of the functional domains of the ADA2 protein and the location of the ADA2 variants identified in our cohort (labeled ‘*’), in previous studies (labeled ‘∼’) and in literature (labeled ‘§’).

Five out of ten patients presented with (muco)cutaneous manifestations (Supplemental Table. 4): two with livedo, one with Raynaud phenomenon and two with non-specific cutaneous vasculopathy, including chilblain-like lesions (Supplemental Figure. 1B). Neurological manifestations were observed in three patients: two patients experienced ischemic strokes and brain MRI of P6 revealed white matter lesions. Representative images are shown in Supplemental Figure. 1A and 1C. Immunological and/or hematological manifestations occurred in four patients, including hypogammaglobulinemia in three patients, an insufficient pneumococcal antibody response in two patients, neutropenia and thrombocytopenia in P3 and a deep venous thrombosis with pulmonary embolism in P4. Increased infectious susceptibility to viral and bacterial infections was noted in three patients. Gastro-intestinal manifestations were present in three patients. P3 experienced abdominal pain, chronic dyspepsia, nodular regenerative hyperplasia and portal hypertension, P9 had abdominal pain and P10 presented with hematemesis. Musculoskeletal symptoms were observed in two patients, both of whom had arthritis, with P10 also experiencing tendinitis. Pericarditis was documented in P10. P7 was the only treated with TNF inhibition (TNFi).

Whole exome sequencing revealed four distinct heterozygous variants in these patients, the p.H424N variant in P1 and P2, the p.G47V variant in P3-5 and P8, the p.R169Q variant in P6 and P7 and the p.G47R variant in P10. Sanger sequencing of gDNA and cDNA confirmed the pathogenic variants identified via whole exome sequencing. No additional pathogenic variants in *ADA2* were identified. Genetic intolerance scores for *ADA2* support ADA2’s role as a recessive gene with limited tolerance for variation, but they do not strongly suggest a dominant pathogenic role in heterozygous mutations (Supplemental Table. 7) (21).

To study the potential effect of heterozygous pathogenic variants in ADA2, we examined the four variants, resulting in single amino acid substitutions in different domains of the ADA2 protein, discovered in P1-P10, along with the heterozygous variants reported in the literature (10, 17–19). The variants G47A and G47W were also assessed, as these proven pathogenic variants are located at the same amino acid position as the G47V and G47R found in P3-5, P8 and P10, respectively (Figure. 1B). Proven pathogenic variants T360A and N370K were also included (16).

### G47V, R169Q, H424N and Y453C ADA2 variants affect secretion of wild-type ADA2 protein

We assessed the effect of ADA2 missense variants on wild-type (WT) ADA2 protein expression, secretion and enzymatic activity. Therefore, we performed transient transfection of each *ADA2* variant alone (homozygous), as well as transient co-transfection of each *ADA2* mutant together with WT *ADA2* to mimic the carrier status (heterozygous) (10, 22). Western blotting of cell lysates on denaturating gels showed that in homozygous condition, ADA2 intracellular protein expression levels of most studied ADA2 variants were comparable to WT100% ADA2, except for G47A, G47R, G47V and Y453C where there was reduction of 20% compared to WT100% (Supplemental Figure. 2A and B, Supplemental Figure. 3A and B, Supplemental Figure. 4A and B). When we assessed ADA2 secretion in homozygous ADA2 variant conditions, no residual ADA2 secretion was observed for the R169Q and Y453C variant (Supplemental Figure. 2A and C). This is in line with the observations by Cheng et al. (23). Compared to WT100%, the secretion of variant H424N was reduced by 80% (Supplemental Figure. 2A and C). A decrease in secretion of around 50% was observed in G47A and G47R when compared to WT100%. The decrease in secretion of variant G47W was more pronounced, with a reduction of 70%. Variant G47V exhibited nearly absent secretion (residual secretion around 10%) (Supplemental Figure. 2A and C). Moreover, variants E328K, T360A and N370K showed a reduction of 30%, 20% and 28% in secretion compared to WT100%, respectively (Supplemental Figure. 2A and C, Supplemental Figure. 4A and C). Variant F355L exhibited normal protein secretion compared to WT100% (Supplemental Figure. 3A and C).

When ADA2 variants were transfected in a heterozygous setup with WT ADA2, we observed a decrease in secretion across all variants except for E328K, F355L, T360A and N370K, when compared to WT50% (Supplemental Figure. 2A and D, Supplemental Figure. 3A and D, Supplemental Figure. 4A and D). While the secretion of ADA2 was only reduced by around 10% for variants G47R, G47V and Y453C, and 20% for G47A and G47W, the reduction below WT50% was more pronounced for R169Q and H424N with a reduction of 70% and 35% respectively (Supplemental Figure. 2A and D). Taken together, our data suggest a dominant negative effect of ADA2 variants R169Q and H424N on WT ADA2 secretion.

Since ADA2 dimer formation is required for ADA2 deaminase activity (24), we next assessed the secreted ADA2 dimer formation of each variant in both homozygous and heterozygous conditions in non-denaturating gels. In homozygous setting, F355L shows normal dimer secretion in homozygous conditions (Supplemental Figure. 5A and B). Variant G47A, G47R and G47W show a residual ADA2 dimer secretion of around 48%, 53% and 9% respectively compared to WT100% (Figure 2A and B).

**Figure. 2.**
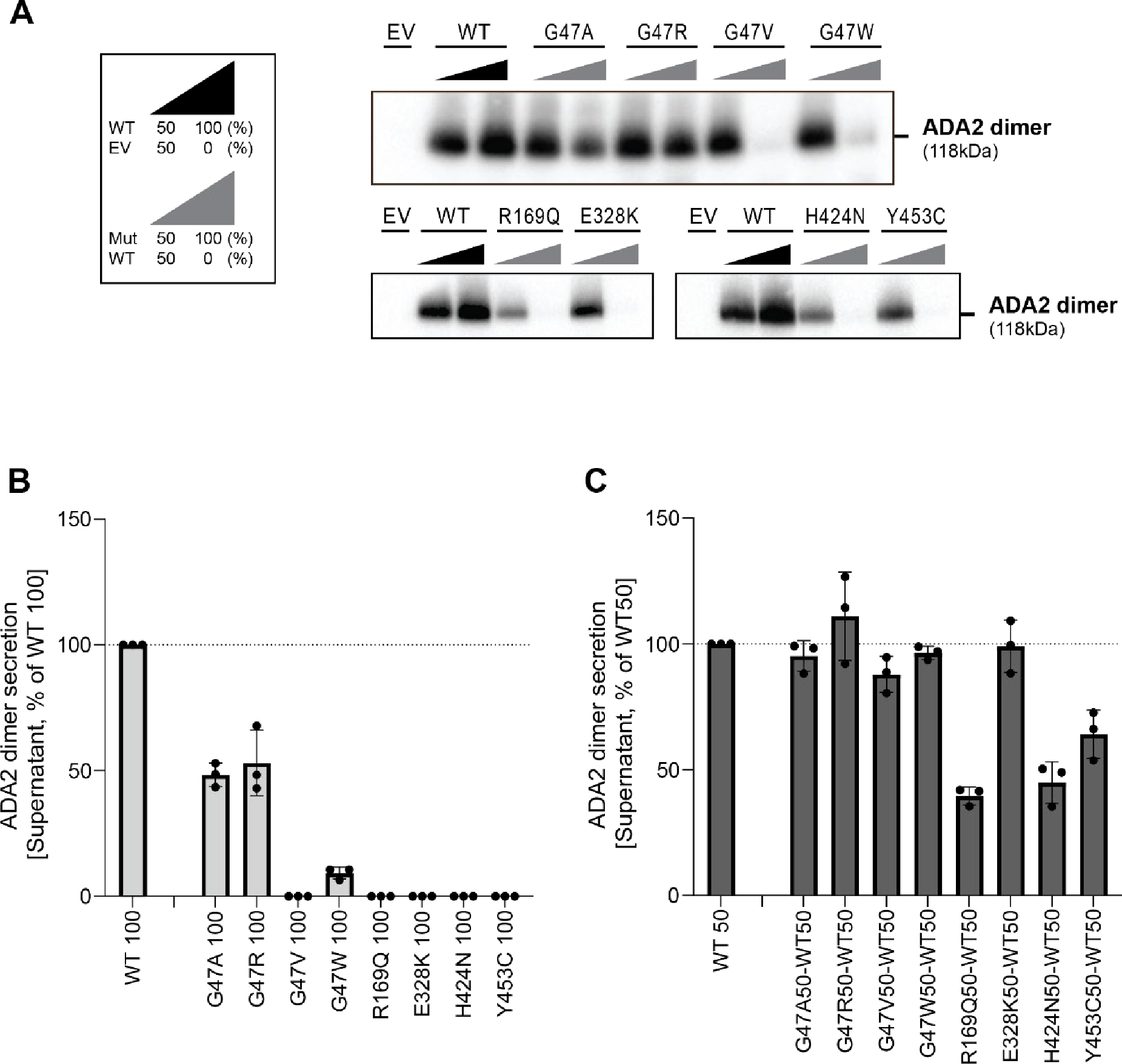
Secretion of ADA2 dimers in homozygous or heterozygous state on non-denaturating gel. **A**. ADA2 dimer secretion of HEK293T cells transfected with WT and/or ADA2 variants. Cells and supernatant were collected 48h after transfection. **B.** Quantification of ADA2 secretion in Supernatant of transfected HEK293T cells with wild-type ADA2 or ADA2 variants in homozygous conditions. Bar graphs represent percentage of ADA2 protein secretion relative to wild-type ADA2 100%. **C.** Quantification of ADA2 secretion of co-transfected HEK293T cells of ADA2 variants together with wild-type ADA2 in heterozygous conditions. Bar graphs represent percentage of ADA2 secretion relative to wild-type ADA2 50%. experiments. **A-C.** Image shown represents 3 independent experiments.

However, no ADA2 dimers were detected in the supernatant of the homozygous G47V, R169Q, E328K, H424N and Y453C transfection setting (Figure. 2A and B). More interestingly, expression of variants T360A and N370K led mostly to secretion of monomeric ADA2 (Supplemental Figure. 6A and B).

In heterozygous setup (WT/M) G47A, G47R, G47W, E328K, T360A and F355L ADA2 variants result in levels of dimer secretion comparable to WT50% (Figure. 2A and C, Supplemental Figure. 4A and C). A small reduction in dimer secretion of 10% was observed in WT/G47V and WT/N370K (Figure. 2A and C, Supplemental Figure. 4A and C). However, R169Q, H424N and Y453C showed a 60%, 50% and 35% decrease respectively in ADA2 dimer secretion when compared to WT50% (Figure. 2A and C). This observation further supports the hypothesis that R169Q, H424N and Y453C exert a dominant negative effect on WT ADA2.

### Enzymatic activity of WT ADA2 is affected in a dominant negative manner by G47A, G47R, G47V, R169Q, E328K, H424N and Y453C ADA2 variants in overexpression

Next, the ADA2 enzymatic activity of each ADA2 variant was evaluated. We observed normal ADA2 enzymatic activity for variant F355L (Supplemental Figure. 7). In the homozygous conditions, a reduction of intracellular ADA2 enzymatic adenosine deaminase activity was observed in all other *ADA2* variants studied, when compared to 100% WT ADA2 (Figure. 3A, Supplemental Figure. 8A). For G47A, G47R, G47V and R169Q the residual enzymatic activity was around 25% compared to WT100%. Variant T360A and N370K show a residual enzymatic activity of 35% and 20% respectively. This reduction was even more pronounced in variants G47W, E328K, H424N and Y453C, where the residual enzymatic activity was only 8%, 7%, 11% and 7,5% respectively (Figure. 3A, Supplemental Figure. 8A). When ADA2 activity of secreted homozygously expressed *ADA2* variants was examined, G47A,G47R, T360A and N370K showed 25%, 6%, 8% and 7% residual enzymatic activity respectively (Figure. 3B), Supplemental Figure. 8B). However, G47V, G47W, R169Q, E328K, H424N and Y453C exhibited nearly absent enzymatic activity.

**Figure 3.**
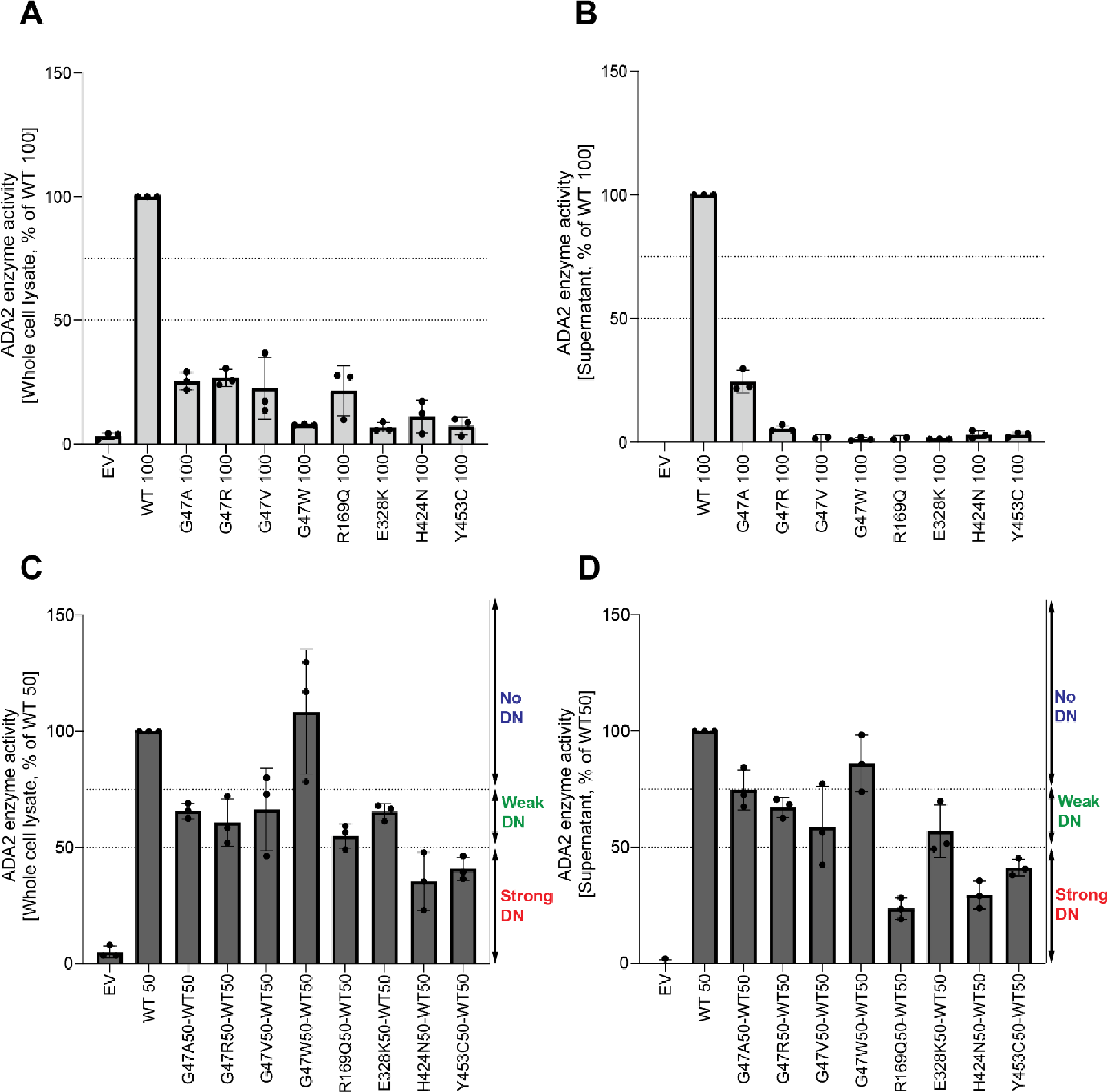
Adenosine deaminase activity of ADA2 variants in homozygous or heterozygous state. **A.** Adenosine deaminase activity in whole cell lysate of HEK293T transfected cells with WT and ADA2 variants in homozygous conditions. Bar graphs represent the percentage of enzymatic activity relative to wild-type ADA2 100%. **B.** Adenosine deaminase activity in supernatant of HEK293T transfected cells with WT and ADA2 variants in homozygous conditions. Bar graphs represent the percentage of enzymatic activity relative to wild-type ADA2 100%. **C.** Adenosine deaminase activity whole cell lysate of HEK293T transfected cells with WT and/or ADA2 variants in heterozygous conditions. Bar graphs represent the percentage of enzymatic activity relative to wild-type ADA2 50%.. **D.** Adenosine deaminase activity in supernatant of HEK293T transfected cells with WT and/or ADA2 variants. Bar graphs represent the percentage of enzymatic activity relative to wild-type ADA2 50%.. **A-D.** Data represents mean ± SD from 3 independent experiments.

In heterozygous conditions, variants G47W, T360A and N370K did not affect the enzymatic activity of WT ADA2 (Figure. 3C and D, Supplemental Figure. 8C and D) unlike G47A, G47R, G47V, R169Q, E328K, H424N and Y453C which resulted in enzymatic activity levels below that of the 50% ADA2 WT gene product, suggesting a dominant negative effect. Based on the effect on WT ADA2 enzymatic activity, we made a distinction between absent dominant negative effect (enzymatic activity >75% of WT50%), a weak dominant negative effect (enzymatic activity 50-75% of WT50) and a strong dominant negative effect (enzymatic activity 0-50% of WT50) as described before (16, 25).

Variants G47A, G47R, G47V and E328K exert a weak dominant negative effect on the enzymatic activity of WT ADA2 both intracellularly as assessed in the whole cell lysate and extracellularly as secreted ADA2 in the supernatant, with a residual enzymatic activity below 75% (Figure. 3C and D). The weak dominant negative effect is most pronounced in the supernatant of variant G47V, with a residual activity of 58% compared to WT50. More interestingly, variant R169Q exhibits a weak dominant negative effect on intracellular ADA2, however it exerts a strong dominant negative effect on the enzymatic activity of secreted WT ADA2 with a residual activity of only 23% (Figure. 3C and D). This is in line with the decrease in dimer secretion observed in the heterozygous R169Q condition. Furthermore, variant H424N has a strong dominant negative effect on both intracellular as well secreted WT ADA2 enzymatic activity (Figure. 3C and D). This suggests that, in addition to hindrance of ADA2 secretion (Figure. 2), there is also an intrinsic inhibition of the WT ADA2 activity. Lastly, variant Y453C exhibits a strong dominant negative effect with a residual activity of only 41% compared to WT50 both intracellularly in the whole cell lysate as well as extracellularly on secreted ADA2 (Figure. 3C and D).

### Prediction model of ADA2 dimer formation in the presence of ADA2 variants

Molecular dynamics (MD) simulations indicate the contributions of amino acid residues to the stability and conformational dynamics of the complex. To investigate these dynamics within the ADA2 complex, three independent 100 ns MD simulations for both the monomeric WT and mutated ADA2 were performed. After completing the simulations, the B-factors for each individual monomer, which serve as a measure of atomic fluctuation within the structure, were calculated. These B-factors were averaged across all monomers and normalized by comparing them to those obtained from the WT simulations, allowing for a relative assessment of the fluctuation.

All variants except Y453C result in increased fluctuation in their immediate vicinity and thereby demonstrate increased structural instability (Figure. 4A). Notably, the variants at residues G47 and H424 exhibit heightened fluctuation, particularly in regions corresponding to the dimer interface. This increase in fluctuation may explain the observed decrease in deaminase activity when these variants occur as it may prevent correct folding, leading to aggregation or the formation of the dimer interface. The pathogenic variant R169Q also exhibited heightened fluctuation; however, given its distance from the interface, or the active site, this results in disrupted folding/stability. In the WT structure, R169 stabilized the fold with a salt bridge with residue D179, hydrogen bonds with residue T129 and aromatic stacking between the guanidinium group of residue R169 and the aromatic ring of residue Y130 (Figure. 4B). All three are disrupted upon mutation. This impairment could be due to the change from a positively charged side chain to an uncharged, shorter side chain, and thus losing interactions with its neighboring residues. Similarly, variant E328K, which is located in the vicinity of the active site will lead to a disruption of a three strong hydrogen bonds with residues N370, G358 and E359, the latter being essential for catalytic activity (24), leading to loss of structural stability as well as catalytic activity (Figure. 4C). The variant Y453C showed a slight decrease in fluctuation in its immediate vicinity (Figure. 4A). However, due to its distance from the active site or dimerization interface, this is not expected to have an effect on either. In the WT structure, Y453 stabilizes the fold with a hydrogen bond with E94, which is lost upon mutation (Figure. 4D). This mutation to a smaller sidechain with loss of interactions is expected to disrupt the folding.

**Figure 4.**
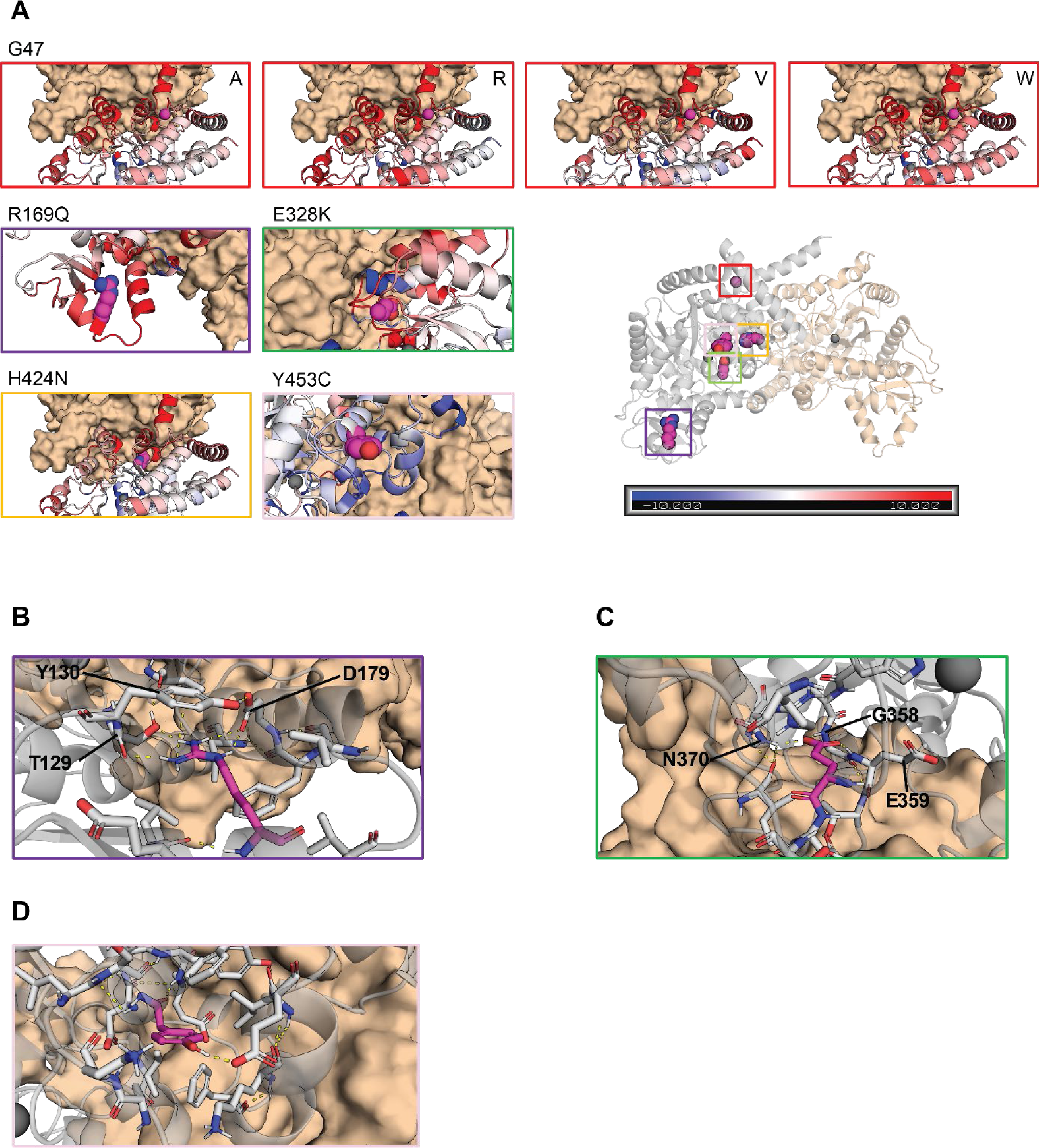
Molecular modeling of ADA2 variants G47A, G47R, G47V, G47W, R169Q, E328K, H424N and Y453C. **A**. Comparative plots of the different mutations which displays the variability of B-factor values along the protein backbone compared to the wild target. Blue backbones represent lower fluctuations compared to the wild target, red backbones represent higher fluctuations. **B.** Zoom-in of residue R169 in the wild target protein. R169 is shown in magenta. Polar and aromatic interactions are shown as dashed lines. **C.** Zoom-in of residue E328 in the wild target protein. E328 is shown in magenta. Polar interactions are shown as dashed lines. **D.** Zoom-in of residue Y453 in the wild target protein. Y453 is shown in magenta. Polar interactions are shown as dashed lines.

### Serum ADA2 enzyme activity of diseased heterozygous carriers of pathogenic ADA2 variants is in the carrier range

To assess whether the dominant negative effect of the indicated variants could be discriminated from the patients’ sera, we measured the ADA2 enzyme activity in the serum of the 10 suspected DADA2 patients. All ADA2 enzyme activity levels were in the range of the heterozygous carriers of pathogenic DADA2 variants (Figure. 5A). While the serum ADA2 activity levels of P4, P5 and P7 were in the lower range of DADA2 carriers, we observed that the serum ADA2 activity levels of P1, P2 and P5 were in the higher carrier range (Figure. 5A). Serum ADA2 levels analyzed by Western Blot showed that ADA2 secretion correlated with the serum residual ADA2 enzymatic activity (Figure. 5B-D).

**Figure 5.**
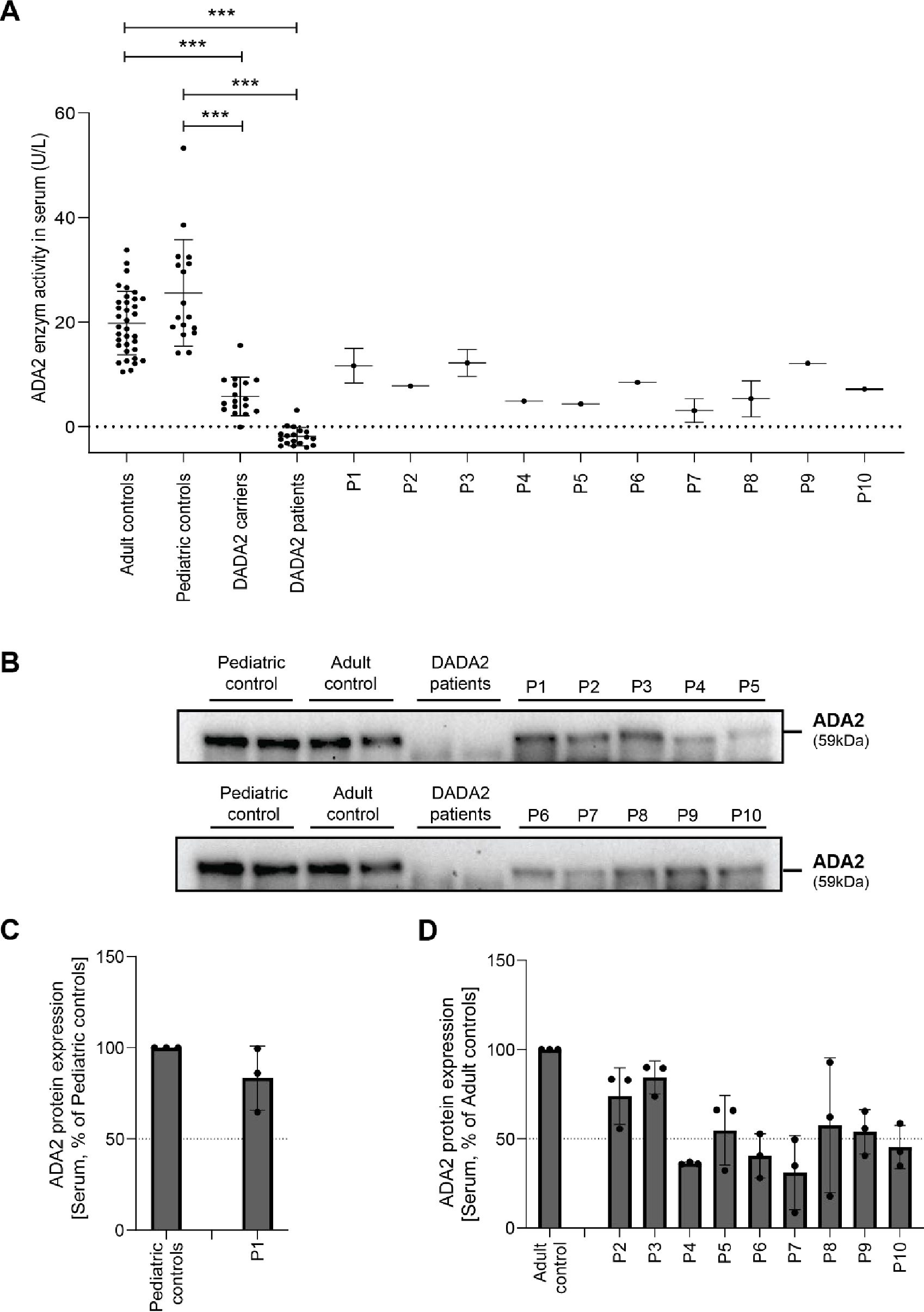
Serum ADA2 enzymatic activity of suspected DADA2 patients. **A**. ADA2 enzyme activity (U/L) measured in serum samples of suspected DADA2 patients (n=10), adult healthy controls (n=35), pediatric healthy controls (n=17), healthy DADA2 carriers (n=19) and DADA2 patients (n=18). Each data point is plotted with mean ± SD. Statistical significance was assessed using the Mann-Whitney U test,*** p<0.0001. **B.** ADA2 protein secretion in Serum samples of pediatric controls, adult controls, DADA2 patients and cohort patients by Western blot. **C-D.** Quantification of ADA2 secretion in Serum samples of pediatric controls, adult controls, DADA2 patients and cohort patients. Bar graphs represent percentage of ADA2 protein secretion relative to Pediatric/adult controls. Each bar represents mean ± SD from 3 independent experiments.

### Population genetics and correlation with phenotypes

Next, we investigated the frequency of the identified dominant negative *ADA2* variants in the general population by looking at the frequency in the overall gnomAD v4.1.0 samples, and by genetic ancestry (Supplemental Table. 8). We found 1,262 alleles for these variants in gnomAD v4. None of the variants were found at the homozygote state implying that about 0.16% of the general population (1,262 out of 807,088 sequenced at this locus) is heterozygote for a dominant negative variant in *ADA2*. The most common variant was R169Q, which has a prevalence above 1 in 1,000 in the European Finnish and non-Finnish genetic ancestry groups. Of note, both p.G47R missense variants were most common in the Middle Eastern and South Asian ancestry groups with an allele frequency close to 1 in 1,000.

Given the relatively high number of individuals in the UK Biobank and from the Finnish population with a R169Q variant, we sought to investigate the clinical impact of these variants in the UK Biobank (26, 27) and in Finngen (28). For the UK Biobank, variant-level pheWAS results were available via two different publicly available resources: (i) Genebass (https://app.genebass.org/) calculated associations for 4,529 phenotypes across 394,841 individuals, and (ii) AZ pheWAS portal (https://azphewas.com/) calculated associations for ∼13K binary and ∼5K continuous phenotypes across ∼500,000 whole-genome sequenced individuals and reported those with a p<0.01. For Finngen, we utilized the data freeze 12 results (available on November 4, 2024), which include association results for 2,502 phenotype endpoints across 500,348 individuals. We focused our analysis of these results to phenotypes known to be relevant to DADA2, and grouped them based on the organs impacted in order to make it easier to compare the results of the three pheWAS analyses, which used different phenotype definitions. For each of the DADA2 phenotypes, we looked whether cases were enriched in R169Q heterozygotes (Table. 1). Several neurological manifestations compatible with DADA2 were more frequent in R169Q heterozygotes, including stroke, sequelae of cerebrovascular disease and headaches. Vascular dementia was also more frequent. We also observed enrichment related to cutaneous vasculopathy such as vasculitis and Raynaud phenomenon, as well as immunological and bone marrow manifestations. Hodgkin lymphoma and myeloid leukemia were also identified during our search which both have been described in DADA2 patients (4, 29–31). We also found DADA2 phenotypes that were not enriched, such as viral warts and cytomegaloviral disease (32, 33).

**Table. 1.**
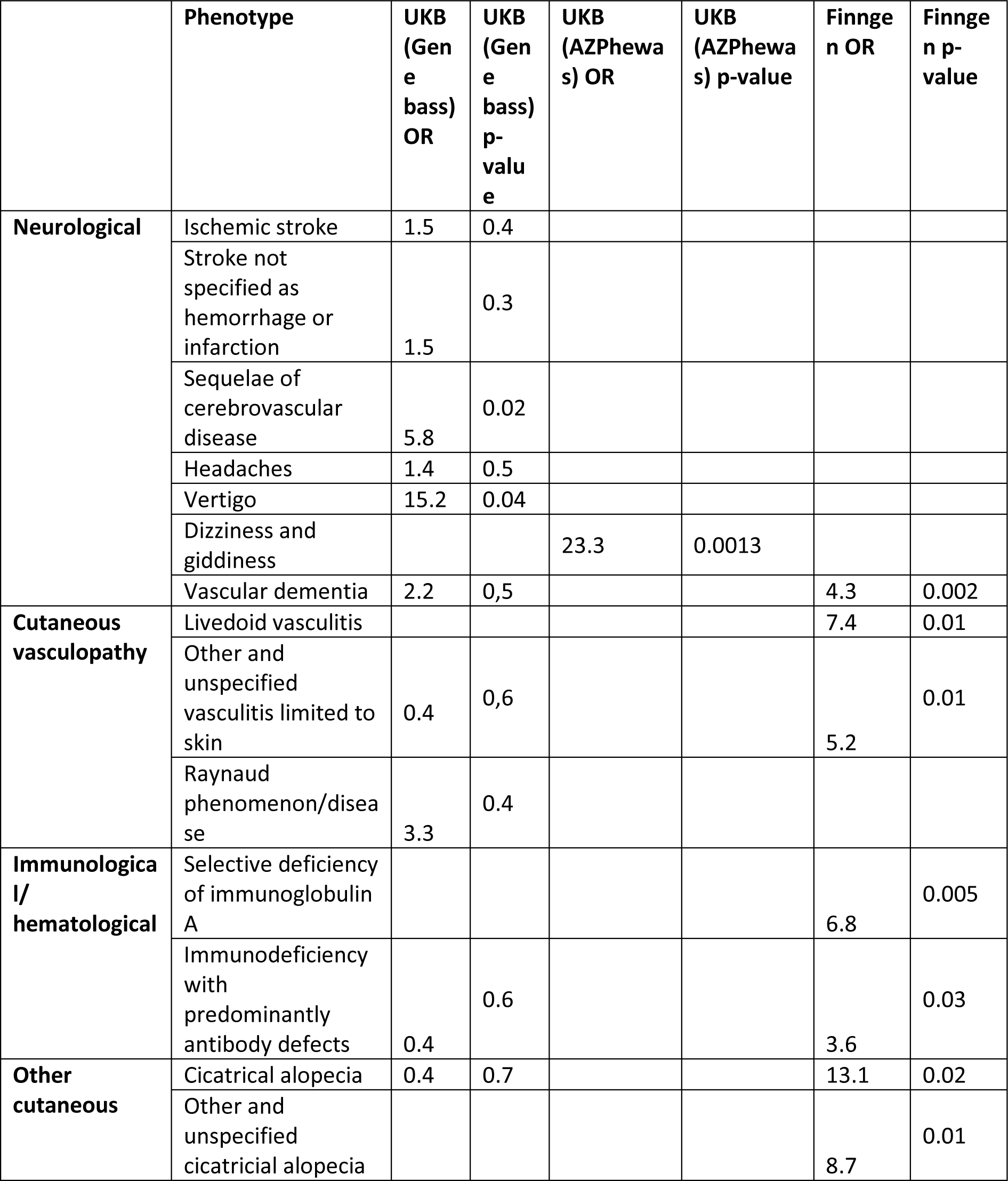
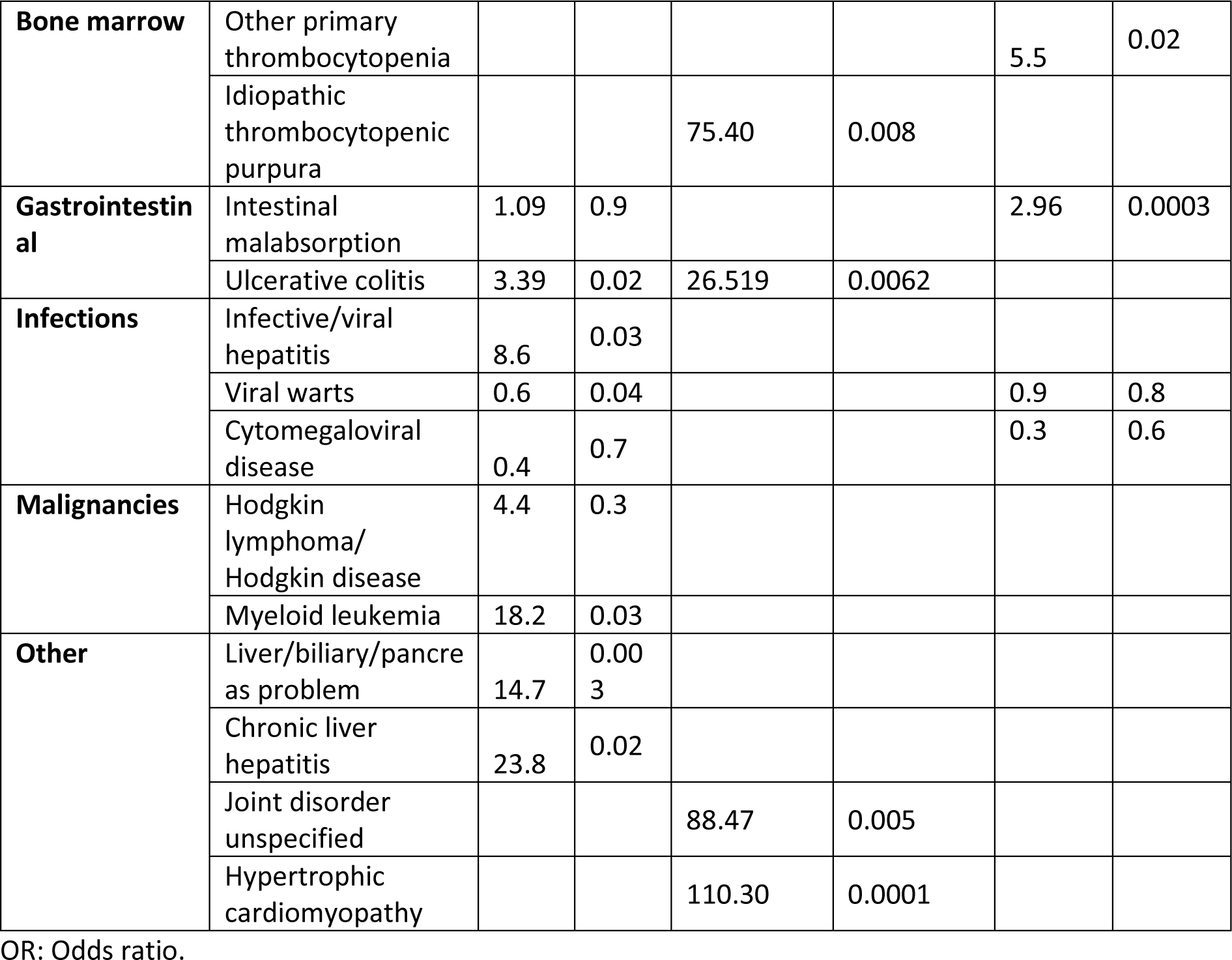
Clinical impact of R169Q in the UK Biobank and Finngen.

To investigate the effect of heterozygous pLOF ADA2 variants in general, we studied the phenotypic associations of collapsed rare pLOF *ADA2* variants in the Bio*Me* BioBank (https://icahn.mssm.edu/research/ipm/programs/biome-biobank/) and the UK Biobank (34). The PheWAS identified several phenotypic associations of *ADA2* pLOFs that align with ADA2 function and clinical presentation of individuals with autosomal recessive DADA2, including diseases of spleen, abnormal results of study function of liver, transient occlusion of retinal artery, abnormal coagulation profile, cranial nerve disorders, paraplegia and diplegia, granulomatous disorders of skin, ulcerative colitis and elevated blood pressure (Supplemental Figure. 9A and B and Supplemental Data File 5 and 6). In conclusion, data from population genetics are supportive of a DADA2 disease state in distinct heterozygous carriers.

## Discussion

Here we report a cohort of heterozygous carriers of pathogenic *ADA2* variants presenting with DADA2 clinical features. In vitro study of the *ADA2* variants identified in this patient cohort revealed that R169Q, H424N and Y453C affect secretion of WT ADA2 protein. Moreover, we demonstrate a dominant negative effect on the enzymatic activity of WT ADA2 by variants G47A, G47R, G47V, R169Q, E328K, H424N and Y453C both intracellularly and extracellularly. Using the HEK293T overexpression model, biochemical modeling and population genetics, we provide proof of principle for the observation that heterozygous carriers of *ADA2* variants can be diseased. In two of the pedigrees, there is an autosomal dominant (AD) pattern of DADA2, albeit with incomplete penetrance. To highlight that the dominant negative effect is variant-specific, we also functionally evaluated ADA2 variants that have no dominant negative effect and that are only pathogenic in homozygous or compound heterozygous state with another pathogenic variant. For variant G47W, the change of a small amino acid (glycine) to a large hydrophobic amino acid (tryptophan) could prevent correct folding which in turn could affect the protein stability and formation of both homodimers as well as heterodimers with WT ADA2. Variants T360A and N370K appeared to be normally secreted, however when secretion of ADA2 dimers was assessed, mainly monomeric ADA2 was detected. This could explain that when present in a homozygous condition, low residual enzymatic activity is observed, since dimer formation is essential for full adenosine deaminase activity (24).

Our study has important clinical implications. First, we provide experimental evidence for the clinical observation of DADA2 disease in heterozygous carriers in the well-established HEK293T overexpression model (16). Based on Jee et al., the carrier frequency of pathogenic *ADA2* variants is estimated to be 1 in 236 when 25% residual enzymatic activity in overexpression is used as cut-off to determine deleterious variants (15). The HEK293T cell overexpression model to study the impact of suspected pathogenic variants has shown that variants associated with disease have variable residual enzyme activity in the supernatant (15, 16). However, although several publications mention disease status in heterozygous carriers (10, 17–19), a potential mechanism has not been explored. Our study shows that different variants exert distinct effects on the WT ADA2 glycoprotein in overexpression. Some variants affect WT ADA2 because of their intrinsic defect in enzymatic activity. Presumably they affect WT ADA2 enzymatic activity when they are assembled with WT ADA2 into a dimer complex. Dimer formation of WT ADA2 with ADA2 variants G47A, G47R, G47V and E328K results in weak dominant negative effect on WT ADA2. When ADA2 variant H424N is built in together with WT ADA2 to form a dimer, the impact on WT ADA2 is more important resulting in a strong dominant negative effect. Other variants affect WT ADA2 by trapping WT ADA2 intracellularly resulting in decreased secretion of WT ADA2 protein and in this way also affects extracellular ADA2 enzymatic activity in a dominant negative manner, as is the case for ADA2 variant R169Q. Beside the in vitro exploration, we studied population databases. Data from PheWAS show that the heterozygous state for pLOF variants in *ADA2* is indeed associated with phenotypes that align with DADA2. When studying the most frequent allele, R169Q, the enriched phenotypes are even more striking, despite the overall low number of cases. Although some phenotypes are soft and subjective like headache, some, like idiopathic thrombopenia and stroke and are more specific and objective. Our data invite therefore to revise the approach to carriers of pathogenic variants for which we describe here a dominant negative effect.

Moreover, our findings stress once again the importance of functional validation of ADA2 variants. Izumo et al. describe a patient with typical DADA2 features yet with serum ADA2 enzymatic activity in carrier range. Two missense variants (F355L and Y453C) were identified (19). However, we show that variant F355L exhibits normal ADA2 enzymatic activity and is normally expressed and secreted. Our data suggest that the Y453C variant is the driving pathogenic variant through dominant negative effect. Likewise another report presents a 22-year-old DADA2 patient who harbors two missense variants, E328K and F355L. Based on our findings, F355L is likely not responsible for the phenotype observed in this patient (35).

Unfortunately, the disease status of carriers cannot be derived in a straightforward way from serum enzyme activity. Indeed, we observed variable ADA2 enzyme activity in the serum of diseased carriers. However, as remarked by Lee and colleagues, the use of the HEK293T overexpression system allows to increase the dynamic range of the measurements. Moreover, current techniques used to measure ADA2 activity in serum are unable to resolve small differences in activity (16). For that reason, the clinical samples of our patients are not able to reflect the findings from the overexpression system. A future more sensitive assay, or a combination of assays or, if feasible, an omics approach may aid in discriminating diseased carriers from patients. In the same way, we previously published that the phenotype and function of lymphocytes from heterozygous carriers were often intermediate to that of healthy donors and ADA2-defcient patients (8). Carmona-Rivera et al. observed increased low density granulocytes in a heterozygous carrier with adult onset polyarteritis nodosa, when studying the role of neutrophils in DADA2 (36).

Interestingly, unlike autosomal recessive (AR) patients harboring homozygous or compound heterozygous pathogenic variants, the patients with heterozygous variants and therefore AD DADA2 seem to present DADA2 manifestations later in life. Indeed, except for P1, P3 and P5, all patients presented after the first decade. We can imagine a situation in which the entire genetic make-up but also the exposure to environmental triggers, like infection or vaccination, tip the balance to the onset of clinically overt inflammation. In addition, by inflammaging, heterozygous carriers of dominant negative *ADA2* variants may naturally evolve towards a DADA2 phenotype (37). Also, except for two patients with a heterozygous R169Q variant presenting with hypogammaglobulinemia and lymphopenia, no cytopenia was observed and most clinical manifestations consisted of vasculitis. This is in line with the proposition by Lee et al. where the lowest residual ADA2 enzyme activities are linked to bone marrow phenotype and the higher residual ADA2 enzyme activities are linked to vasculitis or vasculopathy (16). Of course, it cannot be excluded that modifier genes are at play or that a second hit in a hitherto undefined gene causes the phenotype in these heterozygous carriers, as is the case in a recent paper by Ahn et al (38). Finally, there remains the possibility that monoallelic expression plays a role, although ADA2 is not listed amongst the genes exhibiting this expression pattern (39, 40). The same hypotheses can be put forward to explain the incomplete penetrance of the phenotype in the heterozygous carriers of the dominant negative variants.

Our findings also have important implications for the choice of hematopoietic stem cell donors. Until now, there is debate as to whether procedures with heterozygous carrier donors have worse outcome. However, it can be considered safer to select a matched unrelated donor in cases of siblings carrying monoallelic mutations in ADA2 (13, 41). Our data show that the decision to opt for a matched carrier of a heterozygous variant as donor may depend on the specific variant, highlighting the essence of studying the pathogenicity of a given variant.

Our investigations are hampered by our incomplete understanding of the pathophysiology of DADA2. It is still unclear what the physiological function of ADA2 is and whether the intracellular or extracellular fraction is more important. Until now diagnosis of DADA2 has consisted of identification of two known pathogenic variants or variants of uncertain significance combined with the detection of significantly reduced or absent ADA2 activity in the serum or plasma (3, 14). The new data on the lysosomal role of ADA2 further complicate the view on the pathophysiology of this condition and further research is needed to align the prevailing disease models (11). Nevertheless, the finding by Greiner-Tollersrud et al. that the deaminase activity of ADA2 positively correlates with the DNA binding / editing activity stresses the importance of our findings (11).

Our data also imply that the incidence of AD and AR ADA2 deficiency is higher than the published 1 in 222,164 individuals (15). Indeed, the variants we describe as having a dominant negative effect represent a significant fraction of pathogenic variants. Based on a narrative review by Dzhus et al. in which 495 DADA2 patients reported in literature were included, we calculated the frequency of each variant in the DADA2 patient population (42). Variant G47R (found in P10 of our cohort) is the most common, present in 27% of the patients. R169Q was found in 17% of the patients. G47A, G47V, G47W, E328K and Y453C were more rare, only present in 2%, 3%, 1%, 0,6% and 6% respectively (42). As such, the findings of our work are relevant for an important proportion of DADA2 patients’ kindreds. For optimal family counseling, an effort will need to be undertaken to test all variants in heterozygous setting to establish whether they can act in a dominant negative mechanism and cause AD inheritance.

Finally, the question is whether heterozygous patients harboring a single pathogenic *ADA2* variant with a proven dominant negative effect should receive treatment as for AR DADA2. Given the high proportion of patients (three out of ten in our cohort and three out of seven previously reported in literature) with CNS vascular events as well as renal / splenic infarction (one out of seven previously reported in literature), our findings invite for a reconsideration of treating carriers of dominant negative ADA2 variants with TNFi, as per current guidelines for AR ADA2.

## Materials and Methods

### Patient selection and variant inclusion

This study was approved by the Ethics Committee for Research of Leuven University Hospitals (project numbers: S51577, S63077). Patients were selected based either on clinical phenotype fitting DADA2. In other cases, symptomatic parents of DADA2 patients were included. In addition, we studied the variants in previously reported patients with a DADA2 phenotype, harboring each a single deleterious variant (10, 17–19). G47A, G47W, T360A and N370K, proven pathogenic variants in homozygous setting, were also included (16).

### ADA2 sequencing

Whole exome sequencing (WES) was performed in all patients (Genomics Core Leuven, KU Leuven, Belgium). ADA2 mutations were identified by targeted Sanger sequencing on genomic DNA (in P1-3, P5, P6, P8 and P9) and copy DNA (in P1-3, P5-8) (LGC Genomics). Additional non-pathogenic SNPs observed in ADA2 are presented in Supplemental Table. 1. Primers of targeted Sanger sequencing are available upon request.

### Plasmid construction

ADA2 variant constructs were generated by Q5 site-directed mutagenesis (SDM; New England Biolabs) with human wild-type (WT) ADA2 pCMV6 myc-DKK-tagged plasmid (ADA2 transcript 3, NM_001282225 Origene, Cat# RC238645) as backbone. SDM primer sequences designed with NEBaseChanger are provided in Supplemental Table. 2. Plasmids were amplified in heat shock transformed *E.coli* and purification was performed with QIAprep spin Miniprep kit (QIAgen) according to manufacturer’s instructions. Presence of the introduced variants in the plasmids were verified by Sanger sequencing (LGC Genomics).

### Cell culture and transfection

Human embryonic kidney (HEK) 293T cells were seeded at 2,5x10^5^ cells/6-well in 2ml 24h prior to transfection. Cells were transfected with 25ng of WT and 25ng of mutant *ADA2* plasmids for carrier conditions or 50ng with WT or mutant ADA2 plasmids for homozygous conditions, respectively, using Lipofectamine 2000 Transfection Reagent (Thermofisher Scientific) according to manufacturer’s instruction. Medium was changed 24h after transfection. After 48h, cells and supernatant were collected.

### Western Blotting with Denaturating gel

Transfected cells were collected after 48h and whole cell lysate was obtained by lysing cells in 100µl NP-40 buffer (150 mM NaCl, 50 mM Tris-HCl, 1% NP-40, pH 7.4) supplemented with protease inhibitor (Thermo Fisher Scientific). Equivalent amounts of protein were supplemented with 4x Bolt LDS sample buffer (Thermo Fisher Scientific) and 1x Bolt sample reducing agent (Thermo Fisher Scientific). Supernatant from transfected HEK293T cells were diluted 1:7 in 2x Bolt LDS sample buffer and 1x Bolt sample reducing agent (Thermo Fisher Scientific). Equivalent amounts of serum were diluted in in 2x Bolt LDS sample buffer and 1x Bolt sample reducing agent (Thermo Fisher Scientific) Supplemented lysates and supernatant were denaturared at 70°C for 5min prior to separation on a 4-12% Bis-Tris acrylamide gel. SeeBlue™ Plus2 Pre-stained Protein Standard (Thermo Fisher Scientific) was used as protein molecular weight marker. Proteins were transferred on a PVDF membrane (Thermo Fisher Scientific) and blocked with 5% bovine serum albumin (BSA) in Tris Saline. Membranes were subsequently probed with ADA2 primary antibody (Abcam, ab288296, 1/1000) overnight at 4°C. Membranes were washed and incubated with HRP-conjugated secondary goat anti-rabbit (Abcam, ab205718, 1/5000) for 1h at room temperature. Anti B-actin antibody (Sigma, A5441, 1/9000) was used as loading control. Pierce ECL western blotting substrate (Thermo Fisher Scientific) and SuperSignal West Pico PLUS Chemiluminescent substrate (Thermo Fisher Scientific) were used to visualize HRP activity. Chemiluminescent signals were detected with a ChemiDoc XRS + Imaging system (Bio-rad) and Image lab 6.0.1 Software was used for densitometric quantification.

### Western Blotting with Native gel

Intracellular dimer formation and dimer secretion were analysed on NativePAGE™ 4 to 16%, Bis-Tris (Thermo Fisher Scientific). Equivalent amounts of protein were supplemented with 5% NativePAGE G-250 Sample additive (Thermo Fisher Scientific), 4x NativePAGE sample buffer (Thermo Fisher Scientific), and 1x NativePAGE sample buffer (Thermo Fisher Scientific). Supernatant was diluted 1:4 in 5% NativePAGE G-250 Sample additive (Thermo Fisher Scientific), 4x NativePAGE sample buffer (Thermo Fisher Scientific), and 1x NativePAGE sample buffer (Thermo Fisher Scientific). Gel electrophoresis was performed in accordance to manufacturer’s instructions. NativeMark™ Unstained Protein Standard (Thermofisher Scientific) was used as protein molecular weight marker. Proteins were transferred on a PVDF membrane (Thermo Fisher Scientific) in the presence of NuPAGE transfer buffer supplemented with 10% methanol. Protein standard (Thermofisher Scientific) was visualized using ponceau S solution (Abcam). Blocking and antibody probing was performed as described previously.

### Molecular modeling

The crystal structure and mutated structures of the human ADA2 dimer were prepared using 3D protonation and energy minimization with MOE v.2022.2 (*Molecular Operating Environment (MOE)*, 2024.0601 Chemical Computing Group ULC, 910-1010 Sherbrooke St. W., Montreal, QC H3A 2R7, 2024.); however, only the monomeric form was retained for simulations. Prior to performing triple molecular dynamics (MD) simulations with GROMACS v.2022.3 (43), the CHARMM-GUI webserver (44, 45) was utilized to set up the system, incorporating the 4-point rigid water model (OPC)(46), the ff19SB force field for the protein (47), and the 12–6–4 Lennard-Jones potential model for divalent ions (48). Hydrogen mass repartitioning was applied to the complex to enhance simulation stability without affecting the kinetics of the trajectory or conformational sampling (49, 50).

The complexes were placed at the center of a cubic simulation box with a minimum distance of 1 nm from the box boundaries. The system was then solvated with water and neutralized with Cl⁻ and Na⁺ counter ions, followed by energy minimization using the steepest descent method (51). Interactions were calculated using the Verlet cutoff scheme (52) and the Particle Mesh Ewald (PME) coulomb type (53). The LINCS algorithm was employed to constrain bond lengths (54).

Subsequent to energy minimization, the system was equilibrated to 300 K and 1000 kg/m³ using a 100 ps V-rescale thermostat under an NVT ensemble, followed by a 100 ps equilibration under an NPT ensemble with a V-rescale thermostat and C-rescale barostat (55, 56). Positional restraints were applied to the complex during equilibration. The Parrinello-Rahman barostat (57) was utilized for pressure control during the production run.

Figures were made using PyMOL (The PyMOL Molecular Graphics System, Version 3.0 Schrödinger, LLC.).

### Population genetics

For variant p.R169Q, we looked at the three variant-based phenome wide association studies (pheWAS) results as calculated and reported in GeneBass (https://app.genebass.org/) (26), AZ PheWAS https://azphewas.com/) (27) and in Finngen data freeze 12 (https://www.finngen.fi/en/access_results) (28). Genebass is a resource of exome-based association statistics, made available to the public. The dataset encompasses 4,529 phenotypes with gene-based and single-variant testing across 394,841 individuals with exome sequence data from the UK Biobank (26). AZ PheWAS makes use of the UK Biobank 500k WGS (v2) Public (27). We used the subset of the ∼500,000 whole genome sequenced participants released by the UK Biobank that are high quality and predominantly unrelated to evaluate the association between protein-coding variants with ∼13K binary and ∼5K continuous phenotypes using variant-level and gene-level phenome-wide association studies in 6 ancestry groups (European, Askenazi Jewish, Admixed American, African, East Asian, South Asian). A small number of potentially sensitive phenotypes have been excluded. This portal contains the subset of variant-level associations for which p ≤ 0.01 and the subset of gene-level associations for which p ≤ 0.1. Only variants identified in at least 20 samples were included in the variant-level analysis. Please note that no LD pruning has been performed on these associations. The full pheWAS results for p.R169Q are present in Supplemental Data File 1, 2 and 3.

The Bio*Me* BioBank comprises exome sequencing data and electronic health records from 30,813 genetically diverse participants recruited through Mount Sinai Hospital’s primary care clinics. For the UK Biobank analysis, we utilized 200K exome sequences from participants of European ancestry and their electronic health records. Given the low allele frequency of pathogenic ADA2 variants in the population, we collapsed all heterozygous *ADA2* variants predicted to have a loss-of-function effect (pLOF) as determined by LoGoFunc (58) and LOFTEE (59) and performed two gene-based phenome PheWAS in the Bio*Me* BioBank and the UK Biobank (Supplemental Data File 4). We used Firth’s logistic regression for binary phenotypes obtained from ICD-10 codes matching DADA2 phenotypes (Supplemental Table. 3) and mapped to phecodeX (60) and linear regression for quantitative phenotypes that were curated from laboratory measurements and vital signs. All analyses were adjusted for, age, sex and first ten genetic principal components as covariates.

### ADA2 enzyme assay

Adenosine deaminase 2 activity was determined in whole cell lysates and supernatant from HEK293T cells overexpressing ADA2 as well as in human serum. Adenosine deaminase activity was measured in a colorimetric assay adapted from Giusti and Galanti (61). To inhibit ADA1 activity, Erythro-9-(2-hydroxy-3-nonyl) adenine (EHNA) was used. Triplicate measurements were performed for all samples. Enzymatic activity of ADA2 variants overexpressed HEK293T cells was normalized to the activity of WT ADA2.

### Statistical analysis

Quantification of Western blotting and ADA2 enzyme assay data were presented as mean ± standard deviation (SD). Differences between controls and patient populations were statistically assessed using the Mann-Whitney U test. P-values less than 0.05 were considered statistically significant.

### Data availability

Source data are provided in the Supporting Data values file. Additional data can be obtained upon request to the corresponding author.

## Author contributions

IM, LM, AH, and MW. conceptualized and designed this work. IM, LM, BO, AH, AV, AB, YI, MD, VK, SD, MEK, WVE, LE and MW performed the experiments and carried out data acquisition. BO, GB, PDH and RS provided patient samples. IM, LM, AB, SD, MEK, WVE and MW wrote the manuscript. All authors edited the paper.

## Acknowledgments

IM is a Senior Clinical Investigator at the Research Foundation FWO – Flanders, and is supported by the CSL Behring Chair of Primary Immunodeficiencies through KU Leuven, by the FWO Grants G0C8517N, G0B5120N and G0E8420N and by the Jeffrey Modell Foundation. This project has received funding from the European Research Council (ERC) under the European Union’s Horizon 2020 research and innovation programme (grant agreement No. 948959). This work is supported by ERN-RITA. LE was supported by a PhD Fellowship from the Research Foundation FWO – Flanders (grant 11E0123N). LE is a fellow of the BIH Charité Junior Clinician Scientist Program funded by the Charité – Universitätsmedizin Berlin, and the Berlin Institute of Health at Charité (BIH). MEK and YI were funded by the Charles Bronfman Institute for Personalized Medicine at Icahn School of Medicine, Mount Sinai. This work was supported in part through the computational and data resources and staff expertise provided by Scientific Computing and Data at the Icahn School of Medicine at Mount Sinai and supported by the Clinical and Translational Science Awards (CTSA) grant UL1TR004419 from the National Center for Advancing Translational Sciences. This research was conducted using the UK Biobank resource under application number 53074. WVE acknowledges funding of FWO Grant G095522N was used for this research. RS is supported by an FWO senior clinical investigator fellowship (1805523N).

