## Supplemental Data for "Dominant negative *ADA2* mutations cause ADA2 deficiency in heterozygous carriers"

**SUPPLEMENTAL TABLES AND FIGURES**

Supplemental Table. 1 Additional SNPs identified in *ADA2* via targeted Sanger sequencing. Clinical significance were update from gnomAD v4.1.0 (1). MAF, mean allele frequency

| Patient | Variant | Clinical significance | MAF |
|---|---|---|---|
| P1 | c.159C>T, p.(Asn53=) | Benign | 0.495 |
| | c.213G>A, p.(Met71Ile) | / | 0.00000681 |
| | c.1359T>C, p.(Tyr453=) | Benign | 0.307 |
| | c.1386T>C, p.(Ile462=) | Likely Benign | 0.000102 |
| P2 | c.159C>T, p.(Asn53=) | Benign | 0.495 |
| | c.213G>A, p.(Met71Ile) | / | 0.00000681 |
| | c.1386T>C, p.(Ile462=) | Likely Benign | 0.000102 |
| P3 | c.159C>T, p.(Asn53=) | Benign | 0.495 |
| | c.1359T>C, p.(Tyr453=) | Benign | 0.307 |
| P5 | c.159C>T, p.(Asn53=) | Benign | 0.495 |
| P7 | c.159C>T, p.(Asn53=) | Benign | 0.495 |
| P8 | c.159C>T, p.(Asn53=) | Benign | 0.495 |

Supplemental Table. 2 Overview of site-directed mutagenesis primers

| Variant | Forward primer | Reverse primer |
|---|---|---|
| **G47A** | ATGCGGCTGGCGGGGCGGCTG | CATCTTTTCTTTCAACAACAGATGCGC |
| **G47R** | GATGCGGCTGAGGGGGCGGCT | ATCTTTTCTTTCAACAACAGATGCGCCCGTG |
| **G47V** | ATGCGGCTGGTGGGGCGGCTG | CATCTTTTCTTTCAACAACAGATGCGCCCGTG |
| **G47W** | GATGCGGCTGTGGGGGCGGCT | ATCTTTTCTTTCAACAACAGATGCGCCCGTG |
| **R169Q** | GAGGATTATCAGAAGCGGGTG | CAGCAGAATCCACTTGGAAC |
| **E328K** | GGTGGGGCATAAGGACACTGG | AGGTCAAACCCTGCCACC |
| **F355L** | CTTACTTCTTACACGCCGGAG | GCAGCTTAACGCCATCCT |
| **T360A** | CGCCGGAGAAGCAGACTGGCA | TGGAAGAAGTAAGGCAGCTTAAC |
| **N370K** | TAGACAGGAAAATTCTGGATGCTC | TGGAAGTACCCTGCCAGT |
| **H424N** | CTTGAGGAACAACCCTTAGC | TCAGACACCAGTTTCAGC |
| **Y453C** | GGCTTGTCCTGTGATTTCTATG | TTTGGCACCAAACATAGC |

Supplemental Table. 3 Overview of ICD-10 codes

| | ICD-10 code | Name |
|---|---|---|
| **Liver** | K70.1 | Hepatitis (chronic): alcoholic |
| | K71.- | Hepatitis (chronic): drug-induced |
| | K75.3 | Hepatitis (chronic): granulomatous NEC |
| | K75.2 | Hepatitis (chronic): reactive, non-specific |
| | B15 - B19 | Hepatitis (chronic): viral |
| | K73.0 | Chronic persistent hepatitis, not elsewhere classified |
| | K73.1 | Chronic lobular hepatitis, not elsewhere classified |
| | K73.2 | Chronic active hepatitis, not elsewhere classified |
| | K73.8 | Other chronic hepatitis, not elsewhere classified |
| | K73.9 | Chronic hepatitis, unspecified |
| | K74.6 | Other and unspecified cirrhosis of liver |
| | K76.6 | Portal hypertension |
| | K76.8 | Other specified diseases of liver |
| | K76.9 | Liver disease, unspecified |
| **Stroke** | G46.4 | Cerebellar stroke syndrome |
| | G46.5 | Pure motor lacunar syndrome |
| | G46.6 | Pure sensory lacunar syndrome |
| | G46.7 | Other lacunar syndromes |
| | G46.3 | Brain stem stroke syndrome |
| | G45 | Transient cerebral ischaemic attacks and related syndromes |
| | G45.8 | Other transient cerebral ischaemic attacks and related syndromes |
| | G45.9 | Transient cerebral ischaemic attack, unspecified |
| **Other** | D83 | Common variable immunodeficiency |
| | M32 | SLE |
| | L95.0 | Vasculitis |
| | D60 | Acquired pure red cell aplasia |

| | | |
|---|---|---|
| | D60.1 | Transient acquired pure red cell aplasia |
| | D60.8 | Other acquired pure red cell aplasias |
| | D60.9 | Acquired pure red cell aplasia, unspecified |
| | D61.0 | Constitutional aplastic anaemia |
| | D61.3 | Idiopathic aplastic anaemia |
| | D61.8 | Other specific aplastic anaemia |
| | D61.9 | Aplastic anaemia, unspecified |
| | D70 | Agranulocytosis |
| | D69.3 | Idiopathic thrombocytopenic purpura |
| | D69.4 | Other primary thrombocytopenia |
| | D69.5 | Secondary thrombocytopenia |
| | D69.6 | Thrombocytopenia, unspecified |

Supplemental Table. 4 Clinical manifestations of the ten DADA2 carriers from seven unrelated kindreds. *: recurrent verruca vulgaris; **: upper respiratory tract infections necessitating frequent antibiotic therapy. ***: retinal vasculitis, uveitis and vitritis. ****: termed as erythromelalgia.

| **Patient no. (Kindred no.)** | **P1(F1)** | **P2(F1)** | **P3(F2)** | **P4(F2)** | **P5(F3)** | **P6(F4)** | **P7(F5)** | **P8(F5)** | **P9(F6)** | **P10(F7)** | Cumulative N° of patient |
|---|---|---|---|---|---|---|---|---|---|---|---|
| ***ADA2* mutation** | p.H424N/WT | | p.G47V/WT | | | p.R169Q/WT | | p.G47V/WT | p.R169Q/WT | p.G47R/WT | |
| **Manifestations** | | | | | | | | | | | |
| **(Muco)cutaneous** | | **+** | | **+** | **+** | | | **+** | | **+** | **5/10** |
| Livedo | | + | | | | | | + | | | 2/10 |
| Raynaud phenomenon | | | | + | | | | | | | 1/10 |
| Non-specific cutaneous vasculopathic lesions, including chilblain-like lesions | | | | | +**** | | | | | + | 2/10 |
| **Neurological** | **+** | | | | | **+** | | | **+** | | **3/10** |
| Ischemic stroke | + | | | | | | | | + | | 2/10 |
| White matter lesions | | | | | | + | | | | | 1/10 |
| **Immunological/ hematological** | | | **+** | **+** | **+** | **+** | | | | | **4/10** |
| Hypogammaglobulinemia | | | + | | + | + | | | | | 3/10 |
| Insufficient pneumococcal antibody response | | | + | | | + | | | | | 2/10 |
| Neutropenia | | | + | | | | | | | | 1/10 |
| Thrombocytopenia | | | + | | | | | | | | 1/10 |
| Deep venous thrombosis/ pulmonary embolism | | | | +/+ | | | | | | | 1/10 |
| **Infections** | | | **+** | **+** | **+** | | | | | | **3/10** |
| Viral | | | + | | +* | | | | | | 2/10 |
| Bacterial | | | + | | +** | | | | | | 2/10 |
| **Gastro-intestinal** | | | **+** | | | | | | **+** | **+** | **3/10** |
| Abdominal pain | | | + | | | | | | + | | 2/10 |

| | | | | | | | | | | | |
|---|---|---|---|---|---|---|---|---|---|---|---|
| Chronic dyspepsia | | | + | | | | | | | | 1/10 |
| Nodular regenerative hyperplasia | | | + | | | | | | | | 1/10 |
| Portal hypertension | | | + | | | | | | | | 1/10 |
| Hematemesis | | | | | | | | | | + | 1/10 |
| **Muscoloskeletal** | | | | | | | **+** | | | **+** | **2/10** |
| Arthritis | | | | | | | + | | | + | 2/10 |
| Tenditinis | | | | | | | | | | + | 1/10 |
| **Cardiovascular** | | | | | | | | | | **+** | **1/10** |
| Pericarditis | | | | | | | | | | + | 1/10 |
| **Ocular** | | | | | | **+*****| | | | | **1/10** |
| **Treatment** | | | | | | | **+** | | | | **1/10** |
| TNF-inhibitor | | | | | | | + | | | | 1/10 |

Supplemental Table. 5 Immunological blood results including immunological phenotype, immunoglobulin levels and auto-antibodies of patient 3 (P3), patient 5 (P5) and patient 6 (P6). -: negative; /: not applicable; *, prior to IVIG treatment; **, homogenous nuclear pattern, titer 1:80; ***, p-ANCA titer 1:80.

| | | P1 | P3 | | P5 | P6 | P7 | P9 |
|---|---|---|---|---|---|---|---|---|
| | **Ref. value** | | | | | | | |
| **Hemoglobin** | 12.0-16.0 g/L | 13.4 | 14.4 | 14.0 | 13.3 | 15.8 | 15 | / |
| **Platelets** | 150 000-450 000/L | 297 000 | 159 000 | 163 000 | 265 000 | 259 000 | 313000 | / |
| **White blood count** | 4 500-13 000/µL | 8 820 | 5 850 | 5 760 | 4 890 | 10 650 | 9 670 | 6 970 |
| **Neutrophils** | 1 800-8 000/µL | 4 730 | 3 600 | 3 600 | 2 700 | 5 500 | 6 700 | 4 070 |
| **Monocytes** | 600/µL | 600 | 400 | 300 | 300 | 400 | 500 | 690 |
| **Lymphocytes** | 1 000-5 300/µL | 3 290 | 1 802 | 1 800 | 1 700 | 4 500 | 2 100 | 1 920 |
| **T cells (CD3+)** | 800-3 500/µL | 2 740 | 1 297 | 1 466 | 1 404 | 3 412 | 1 592 | 1 425 |
| CD4+ | 400-2 100/µL | 1 722 | 749 | 908 | 891 | 2 611 | 951 | 943 |
| CD8+ | 200-1 200/µL | 822 | 429 | 457 | 434 | 836 | 620 | 362 |
| CD56+ | 4.3–16.2% of CD3+ | 3.2% | 12.7% | 10.4% | 83% | 0.3% | 3.4% | 4.5% |
| HLA-DR+ | 2.3-8.6% of CD3+ | 12.3% | 6.4% | 7.5% | 4% | 11.1% | 15.0% | 12.8% |
| CD27+ CD45RA+ | 40.9-65.7% of CD3+ | 72.5% | 49% | 46.5% | 66.8% | 56.5% | 36.1% | 42.1% |
| CD4+/CD25+CD127 low | 5.0-12.0% of CD4+ | / | 5.6% | 8.9% | 7.2% | 6% | / | / |
| T-cell receptor αβ TcR | 87-99.3% of CD3+ | 94.6% | 88.10% | 90.26% | 94.3% | 98.53% | 99.46% | 92.94% |
| γδ TcR | 3.3-10% of CD3+ | 5.5% | 10.5% | 10.0% | 5.2% | 1.4% | 0.3% | 6.3% |
| CD3+/CD4-CD8- | 4.3–10.7% of CD3+ | 5.2% | 9.7% | 9.0% | 5.6% | 1% | 1.5% | 7.0% |
| **B cells (CD19+)** | 200-600/µL | 394 | 251 | 222 | 209 | 956 | 252 | 209 |
| CD27+ IgM+ IgD+ | 2.6-13.4% of CD19+ | 6.4% | 9.9% | 22.4% | 12% | 9.4% | 25.6% | 7.8% |
| CD27+ IgM- IgD- | 4.0-21.2% of CD19+ | 5.4% | 5.9% | 10.6% | 8.4% | 4.3% | 14.5% | 16.5% |
| CD27- IgM+ IgD+ | 61.6-87.4% of CD19+ | 84.7% | 78.5% | 61.7% | 72.1% | 84.8% | 53.7% | 70.1% |
| **NK cells (CD3-/CD56 and/or CD16+)** | 70-1200/µL | 145 | 191 | 86 | 105 | 100 | 268 | 293 |
| **IgG** | 5.76-12.65 g/L | 11.90 | 5.03* | 12.5 | 10.7 | 4.43* | 17.10 | / |

| | | | | | | | | |
|---|---|---|---|---|---|---|---|---|
| IgG2 | 1.06-6.10 g/L | 1.53 | 1.05* | 4.10 | 3.03 | 0.77* | 5.43 | |
| IgG3 | 0.18-1.63 g/L | 0.78 | 0.33* | 0.33 | 0.27 | 0.18* | 0.36 | |
| **IgA** | 0.81-2.32 g/L | 2.70 | 0.44* | 0.84 | 0.68 | 1.37* | 1.56 | |
| **IgM** | 0.30-1.59 g/L | 0.90 | 1.29* | 1.22 | 0.66 | 0.75* | 1.69 | |
| **Lymphocyte stimulation test** | | | | | | | | |
| Candida-index | ≥ 5.00 | / | 43.17 | / | / | / | / | / |
| Tetanus toxoid-index | ≥ 5.00 | | 5.53 | | / | 132.27 | | / |
| PHA-index | ≥ 5.00 | | 31.04 | | 25.08 | 247.82 | | 44.55 |
| **Pneumococcal antibody response** | Prior to vaccination | | | | | | | |
| Pn type 8 | 0.5 mg/L | / | 2.0 mg/L | / | | 1.4 mg/L | / | / |
| Pn type 9N | 0.7-0.9 mg/L | | 2.3 mg/L | | / | 0.8 mg/L | | |
| Pn type 15B | 1.2-1.9 mg/L | | 4.6 mg/L | | | 1.7 mg/L | | |
| **Auto-antibodies** | | | | | | | | |
| ANA | | | - | - | - | - | positive** | |
| ANCA | | | - | - | - | - | positive*** | |
| Anti-parietal cell antibody - IIF | | | - | | | | | |
| Intrinsic factor antibody | | | - | | | | | |
| Smooth muscle | | | - | | | | | |
| Mitochondria – IIF | | | - | | | | | |
| Cardiac muscle | | | - | | | | | |
| Skeletal muscle | | | - | | | | | |
| Pancreas | | | - | | | | | |
| Adrenal gland | | / | - | | | | | / |
| Glutamic acid decarboxylase 65kDa | < 0.9 kU/L | | < 0.1 kU/L | | | | | |
| Insulin | ≤ 5% | | 1% | | | | | |
| Skin | | | - | | | | | |
| Salivary gland | | | - | | | | | |
| Deamidated gliadin IgG | ≤ 20.0 CU | | < 2.8 CU | | | | | |
| TPO | ≤ 34 IU/mL | | 9 IU/mL | | | | | |
| TSH-receptor antibodies | ≤ 1.0 IU/L | | < 1.0 IU/L | | | | | |
| Liver-kidney-microsome – IIF | | | - | | | | | |

Supplemental Table. 6 Genetic characteristics and in silico prediction of pathogenicity of mutations in *ADA2* identified by whole exome sequencing. Genomic position according to the hg18 (GRCh38) physical position. NM_001282225.1 was used as reference transcript. CADD: Combined Annotation-Dependent Depletion; Polyphen: Polymorphism Phenotyping v2; AF: Allele Frequency, AF was derived using the gnomAD browser; SIFT: Sorting Intolerant From Tolerant (1).

| **Patient** | **P1-2** | **P3-5, P8** | **P6-7, P9** | **P10** |
|---|---|---|---|---|
| **Chromosome** | 22 | | | |
| **Genomic position (substitution)** | g.17181992G>T | g.17209538C>A | g.17207107C>T | g.17209539C>T |
| **Ref SNP cluster ID** | rs1416783635 | rs200930463 | rs77563738 | rs202134424 |
| **cDNA position (substitution)** | c.1270C>A | c.140G>T | c.506G>A | c.139G>A |
| **Protein position substitution** | p.H424N | p.G47V | p.R169Q | p.G47R |
| **Zygosity** | Heterozygous | | | |
| **AF** | 0.0000006196 | 0.000048 | 0.00047 | 0.00006692 |
| **CADD** | 24.8 | 21.5 | 21.4 | 22.9 |
| **MSC** | 3.2 | | | |
| **Polyphen-2** | Probably damaging | | | |
| **SIFT** | Pathogenic supporting | | | |

Supplemental Table. 7 Genetic intolerance scores for ADA2. f parameter: frequency parameter; lofTool: loss-of-function Tool; SIS: Selection Intensity Score; evoTol: Evolutionary Tolerance Score; RVIS: Residual Variation Intolerance Score; pLI: Probability of Loss-of-Function Intolerance; LOEUF: Loss-of-Function Observed/Expected Upper Fraction; CoNeS: Combined Network Score; IEI classification: Inborn Errors of Immunity Classification; IEND classification: Inborn Errors of Neurodevelopment classification; hOMIM classification: Human Online Mendelian Inheritance in Man Classification; IEI mode: Mode of Inheritance for Inborn Errors of Immunity; IEND mode of dominance: Mode of Dominance for Inborn Errors of Neurodevelopment; EOHP/LOIP: Early-Onset/ Late-Onset Immunodeficiency Predisposition; DOMINO: Dominance Inference for Inherited Disease Genes; p(HI): Probability of Haploinsufficiency; SCoNeS: Single Cell Network Score; SCoNeS in leave-one-out: Single Cell Network Score in Leave-One-Out Analysis.

| **Gene** | ***ADA2*** |
|---|---|
| f parameter | 0,56162038 |
| lofTool | 0,161 |
| SIS | 0,41382086 |
| evoTol | 4,7380157 |
| RVIS | -0,10570191 |
| pLI | 8,1396E-08 |
| LOEUF | 0,68448 |
| CoNeS | -0,115379718 |
| IEI classification | IEI AR |
| IEND classification | IEI |
| hOMIM classification | IEI |
| IEI mode | NA |
| IEND mode of dominance | NA |
| EOHP/LOIP | LOIP |
| DOMINO | 0,069576 |
| p(HI) | 0,37 |
| SCoNeS | 0,994 |
| SCoNeS in leave-one-out | 0,992 |

Supplemental Table. 8 Frequency of ADA2 dominant negative variants in the general population from gnomAD v4.1.0 (1) * ENST00000399837.8, ** There are 0 homozygotes in gnomAD for any of these variants

| Variant MANE transcript* protein impact | GRCh38 coordinates | Allele count** in gnomAD v4.1.0 | Allele Number in gnomAD v4.1.0 | Pop max in gnomAD | AF Pop max in gnomAD |
|---|---|---|---|---|---|
| p.G47V | 22-17209538-C-A | 48 | 1,613,586 | Middle Eastern | 0.00033 |
| p.G47A | 22-17209538-C-G | 78 | 1,613,704 | Admixed American | 0.000083 |
| p.G47R | 22-17209539-C-T | 108 | 1,613,980 | Middle Eastern | 0.00082 |
| p.G47R | 22-17209539-C-G | 34 | 1,613,982 | South Asian | 0.00011 |
| p.R169Q | 22-17207107-C-T | 810 | 1,614,176 | European (Finnish) | 0.0018 |
| p.E328K | Not present | Not present | NA | NA | NA |
| p.H424N | 22-17181992-G-T | 1 | 1,613,878 | NA | NA |
| p.Y453C | 22-17181904-T-C | 183 | 1,614,136 | European (non-Finnish) | 0.00014 |

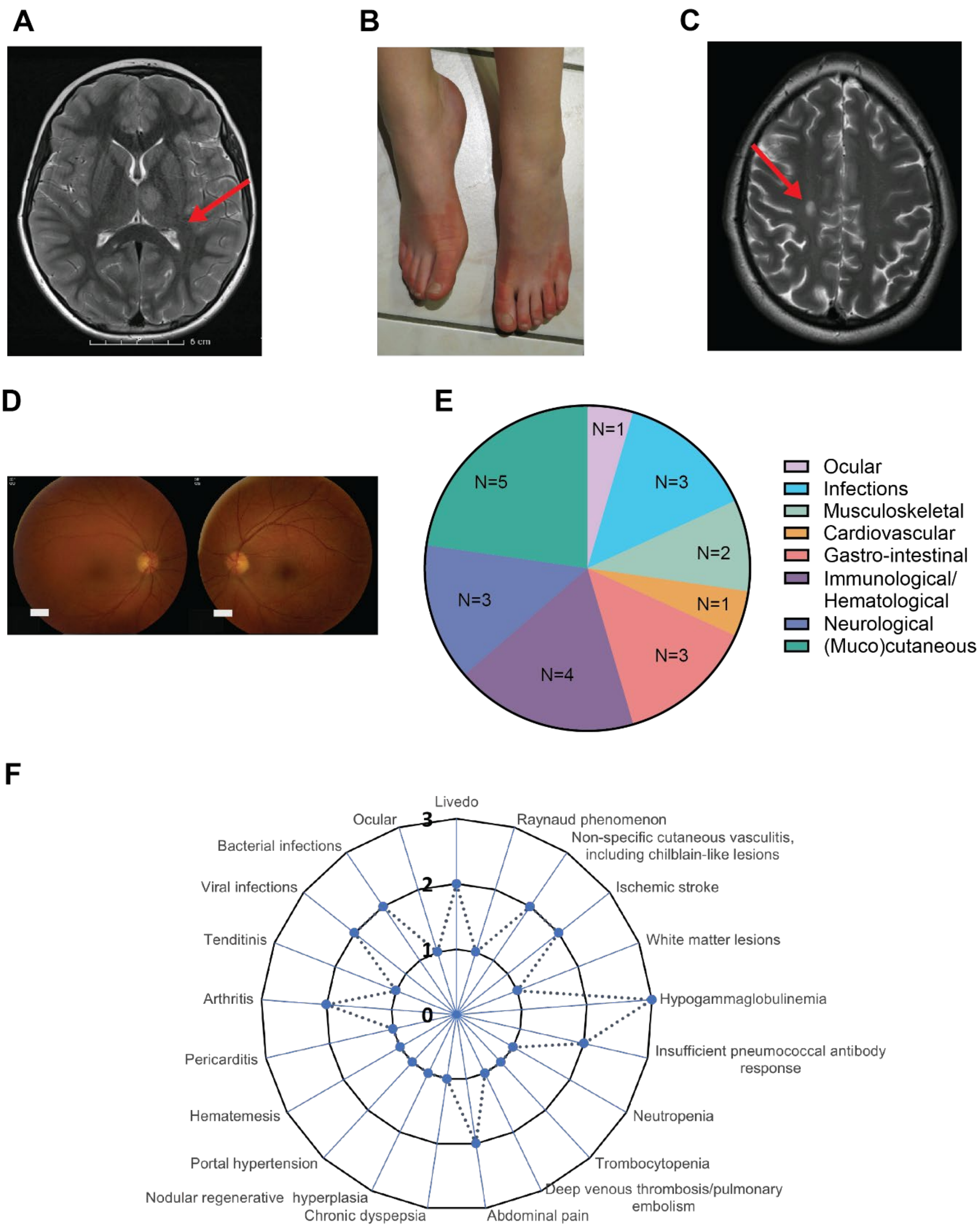


Supplemental Figure 1. Clinical and radiographic findings of the 10 DADA2 carriers.

**A**. Brain MRI of P1 revealing a diffusion restrictive T2-weighted hyperintense lesion with focus anteromedially in the left thalamus, indicating a recent ischemic infarct. **B**. Clinical image of P5 displaying painful purple to red skin discoloration and swelling of the feet. **C.** Brain MRI of P6 showing an oval lesion in the right centrum semiovale, hyperintense on T2 and FLAIR, hypointense on T1, with restricted diffusion and a maximum diameter of 11 mm. **D**. Fundoscopy of P6. The right eye (OD, oculus dexter) shows retinal vasculitis and retinitis with inferiorly located snow ball opacities; left eye (OS, oculus sinister) shows retinitis and vitritis. **E.** Circle diagram illustrating the phenotype distribution by absolute number of affected patients**. F.** Radar graph representing the number of patients affected by various clinical manifestations.

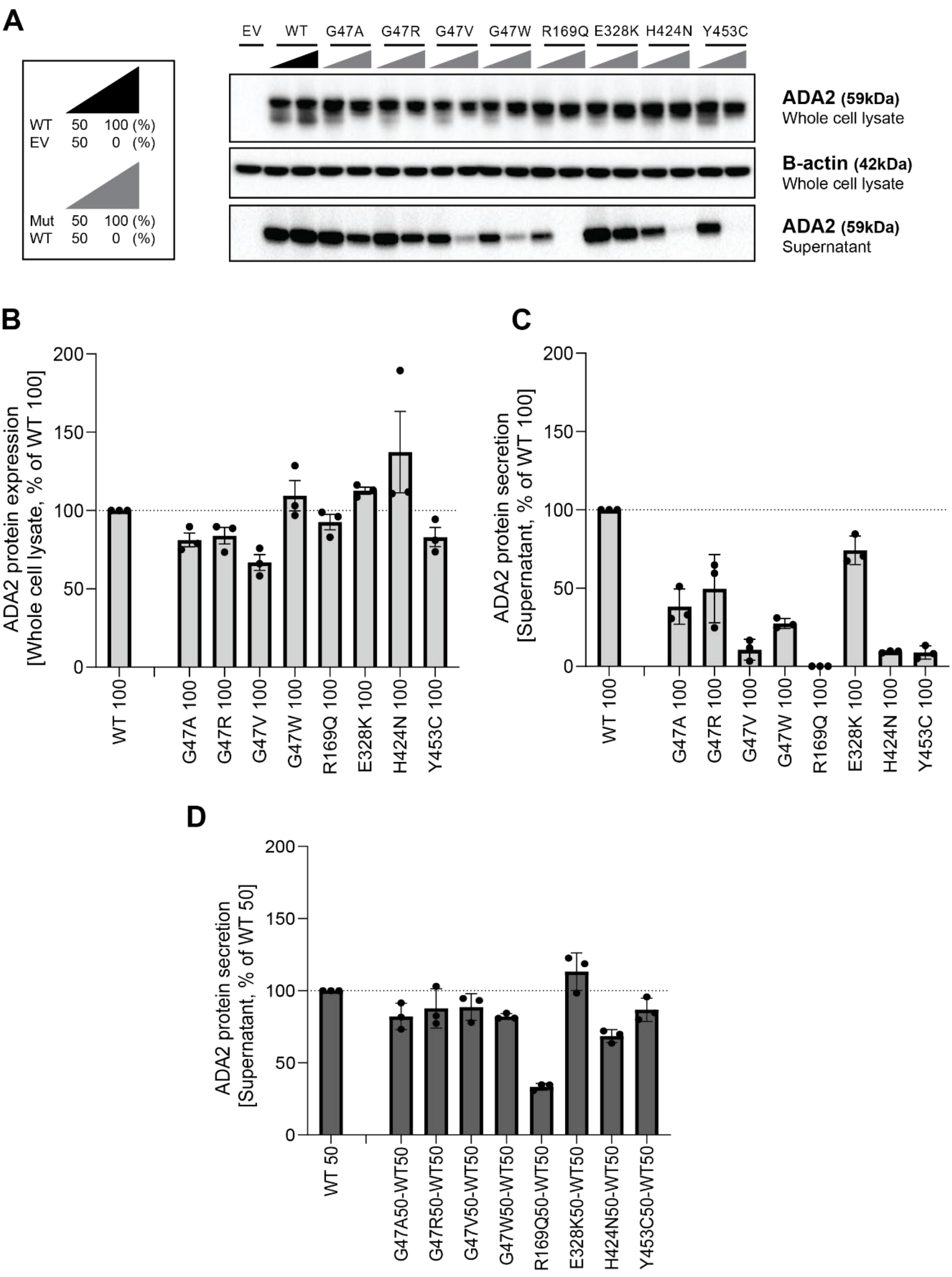


Supplemental Figure 2. ADA2 protein expression and secretion in homogenous and heterozygous state on denaturing gel.

**A.** Immunoblot of whole cell lysate and supernatants of HEK293T cells transfected with different ADA2 variants in homozygous state or together with WT ADA2 (heterozygous state). Cells and supernatant were collected 48h after transfection. Image shown represents 3 independent experiments. Loading control: B-actin. **B.** Quantification of ADA2 protein expression in whole cell lysate of transfected

HEK293T cells with wild-type ADA2 or ADA2 variants in homozygous conditions. Bar graphs represent percentage of ADA2 protein expression relative to wild-type ADA2 100%. **C.** Quantification of ADA2 secretion in supernatant of transfected HEK293T cells with wild-type ADA2 or ADA2 variants in homozygous conditions. Bar graphs represent percentage of ADA2 protein expression relative to wild-type ADA2 100%. **D.** Quantification of ADA2 secretion in supernatant co-transfected HEK293T cells of ADA2 variants together with wild-type in heterozygous conditions. Bar graphs represent percentage of ADA2 secretion relative to wild-type ADA2 50%. **A-D.** Each bar represents mean ± SD from 3 independent experiments.

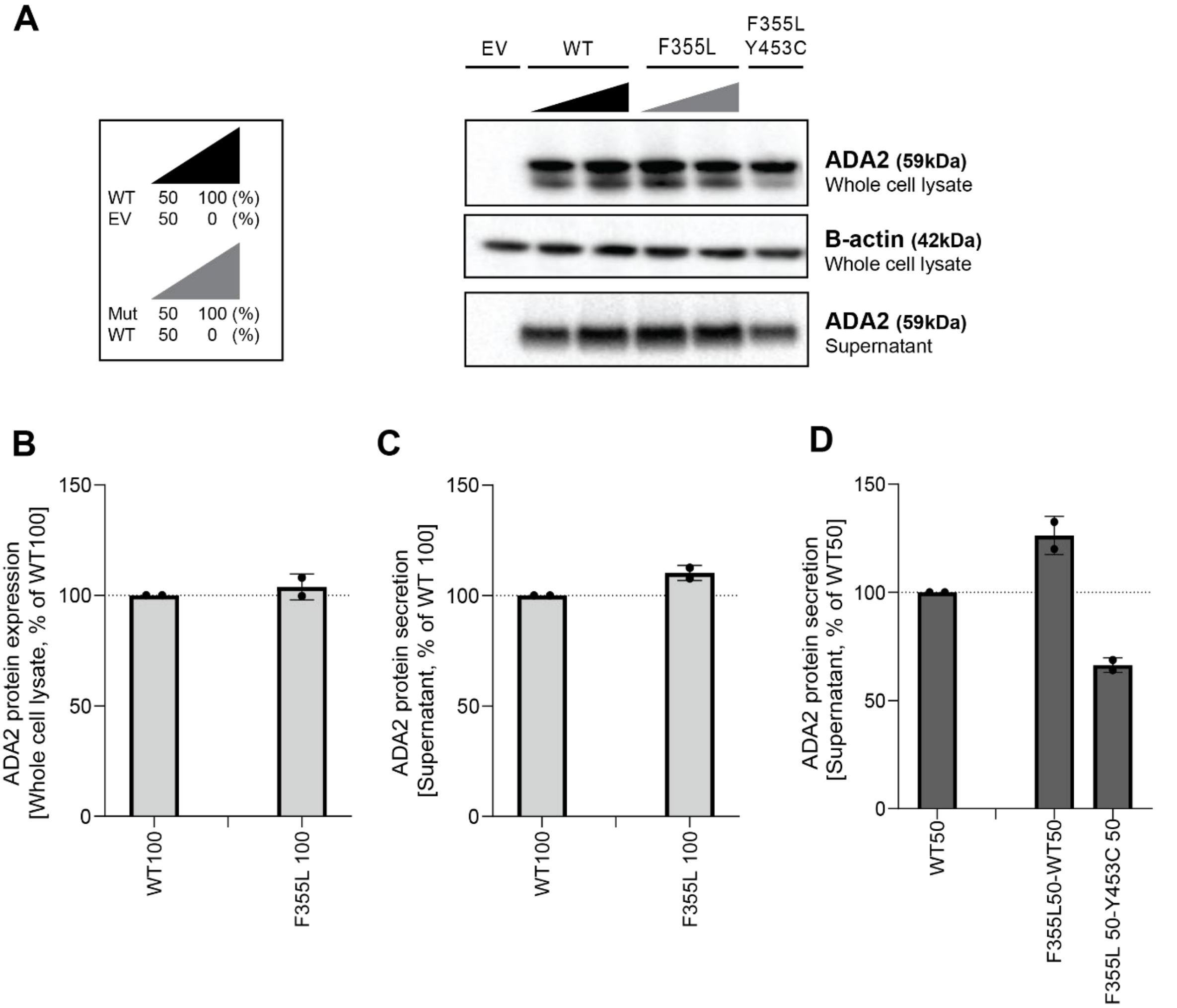


<u>Supplemental Figure 3. ADA2 protein expression and secretion in homogenous and heterozygous state of variant F355L on denaturing gel</u>.

**A.** Immunoblot of whole cell lysate and supernatants of HEK293T cells transfected with ADA2 variant F355L in homozygous state, together with WT ADA2 or with ADA2 variant Y453C in carrier state. Cells and supernatant were collected 48h after transfection. Image shown represents 2 independent experiments. Loading control: B-actin. **B.** Quantification of ADA2 protein expression in whole cell lysate of transfected HEK293T cells with wild-type ADA2 or ADA2 variant F355L in homozygous conditions. Bar graphs represent percentage of ADA2 protein expression relative to wild-type ADA2 100%. **C.** Quantification of ADA2 secretion in supernatant of transfected HEK293T cells with wild-type ADA2 or ADA2 variant F355L in homozygous conditions. Bar graphs represent percentage of ADA2 protein expression relative to wild-type ADA2 100%. **D.** Quantification of ADA2 secretion of co-transfected HEK293T cells of ADA2 variant F355L together with wild-type in heterozygous conditions. Bar graphs represent percentage of ADA2 secretion relative to wild-type ADA2 50%. . **A-D.** Each bar represents mean ± SD from 2 independent experiments.

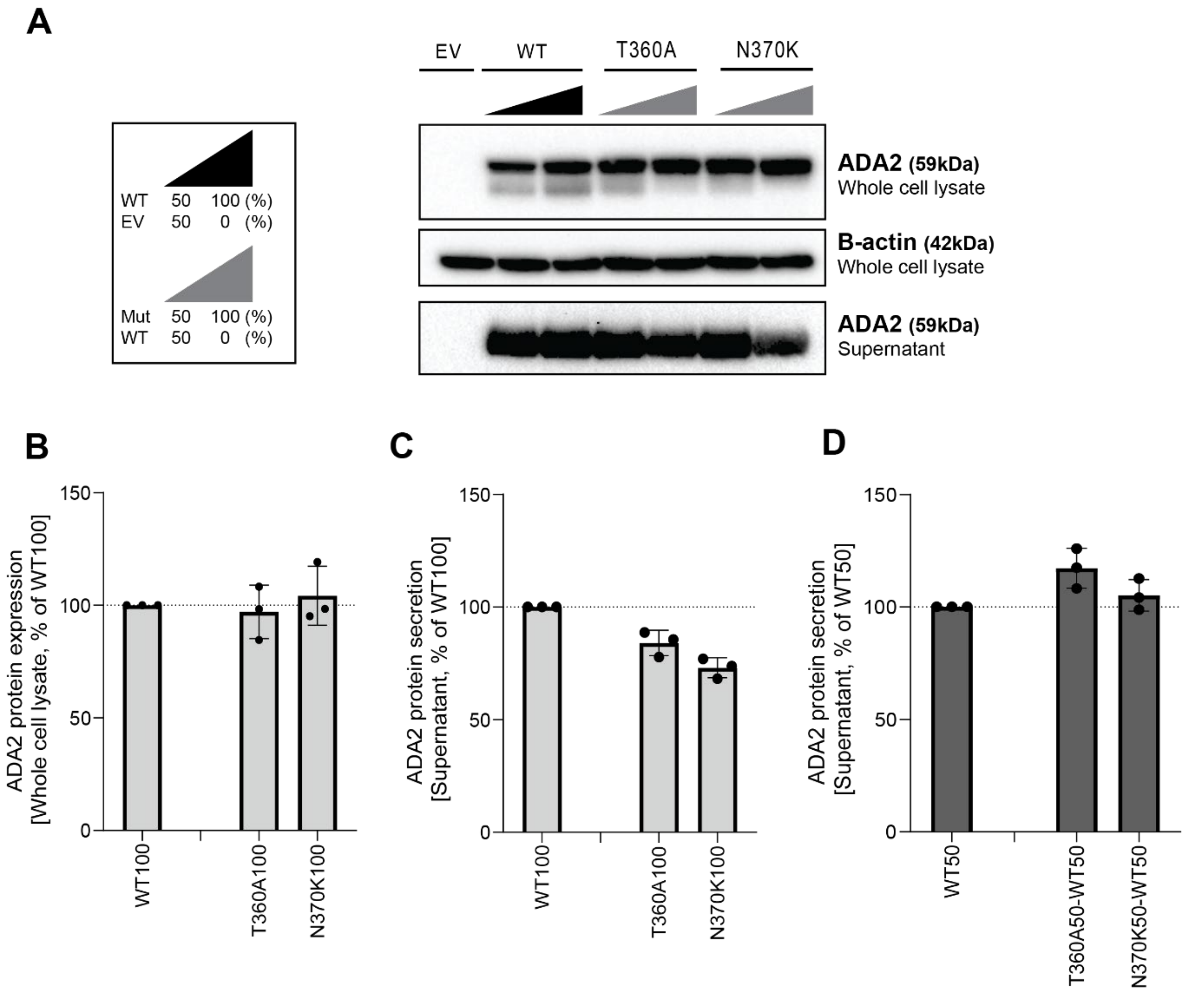


<u>Supplemental Figure 4. ADA2 protein expression and secretion in homogenous and heterozygous state of variants T360A and N370K on denaturing gel</u>.

**A.** Immunoblot of whole cell lysate and supernatants of HEK293T cells transfected with ADA2 variants T360A and N370K in homozygous state or together with WT ADA2. Cells and supernatant were collected 48h after transfection. Image shown represents 3 independent experiments. Loading control: B-actin. **B.** Quantification of ADA2 protein expression in whole cell lysate of transfected HEK293T cells with wild-type ADA2 or ADA2 variants T360A and N370K in homozygous conditions. Bar graphs represent percentage of ADA2 protein expression relative to wild-type ADA2 100%. **C.** Quantification of ADA2 secretion in supernatant of transfected HEK293T cells with wild-type ADA2 or ADA2 variants T360A and N370K in homozygous conditions. Bar graphs represent percentage of ADA2 protein expression relative to wild-type ADA2 100%. **D.** Quantification of ADA2 secretion of co-transfected HEK293T cells of ADA2 variants T360A and N370K together with wild-type in heterozygous conditions Bar graphs represent percentage of ADA2 secretion relative to wild-type ADA2 50%. experiments. **A-D.** Each bar represents mean ± SD from 3 independent experiments.

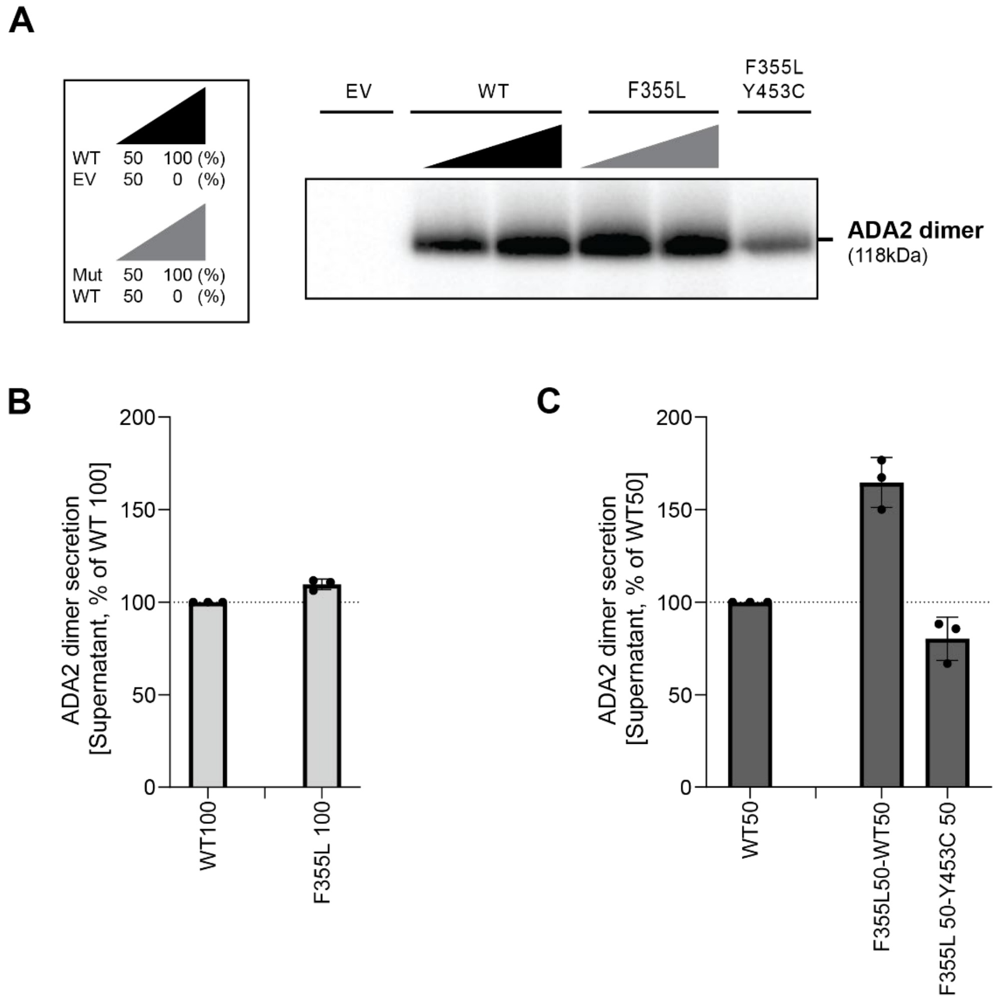


<u>Supplemental Figure 5. Secretion of ADA2 dimers in homozygous or heterozygous state of variant F355L on non-denaturating gel.</u>

**A.** ADA2 dimer secretion of HEK293T cells transfected with WT and/or ADA2 variant F355L. Cells and supernatant were collected 48h after transfection. Image shown represents 3 independent experiments. **B.** Quantification of ADA2 secretion in supernatant of transfected HEK293T cells with wild-type ADA2 or ADA2 variant F355L in homozygous conditions. Bar graphs represent percentage of ADA2 protein secretion relative to wild-type ADA2 100%. **C.** Quantification of ADA2 secretion of co-transfected HEK293T cells of ADA2 variant F355L together with wild-type ADA2 in heterozygous conditions. Bar graphs represent percentage of ADA2 secretion relative to wild-type ADA2 50%. **B-C.** Each bar represents mean ± SD from 3 independent experiments.

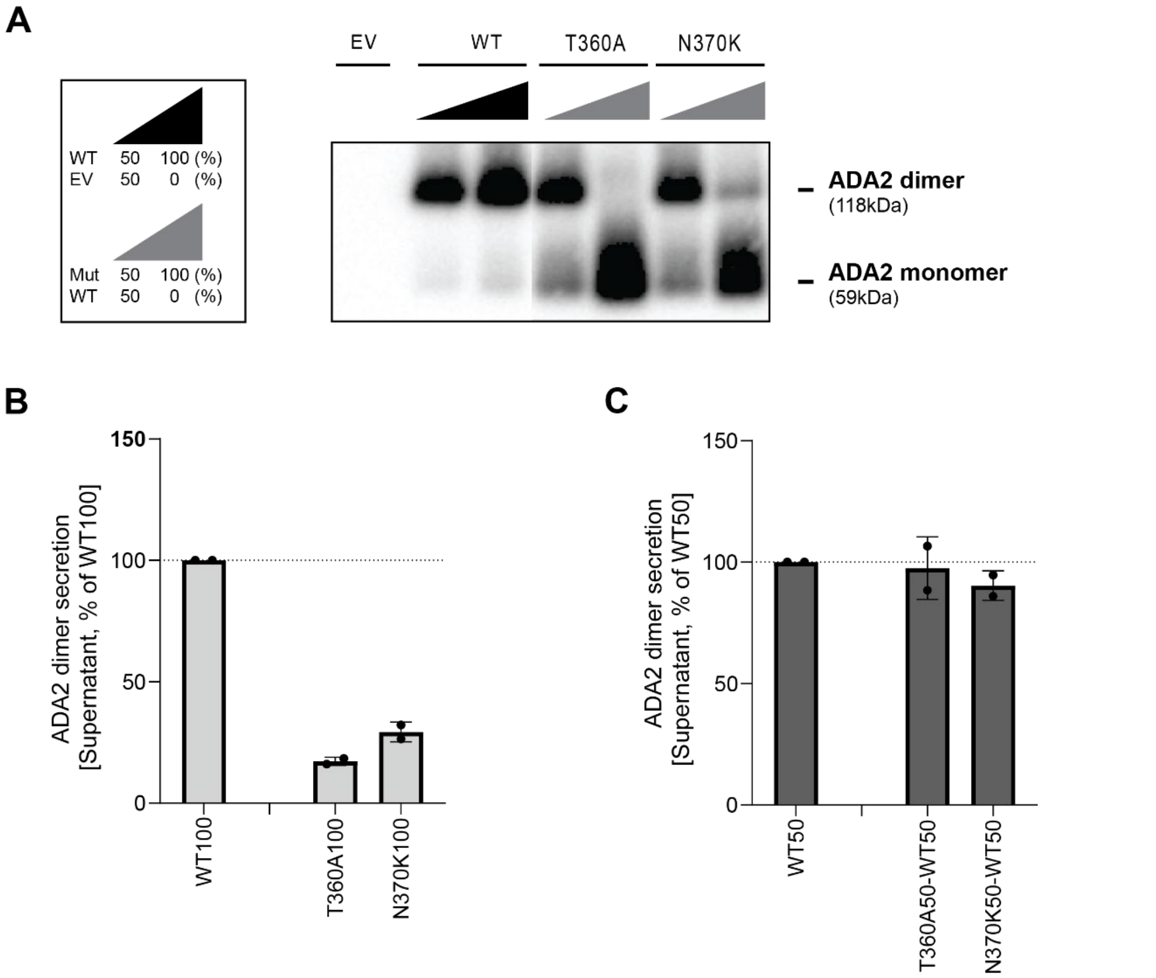


<u>Supplemental Figure 6. Secretion of ADA2 dimers in homozygous or heterozygous state of ADA2 variants T360A and N370K on non-denaturating gel.</u>

**A.** ADA2 dimer secretion of HEK293T cells transfected with WT and/or ADA2 variants T360A and N370K. Cells and supernatant were collected 48h after transfection. Image shown represents 2 independent experiments. **B.** Quantification of ADA2 secretion in supernatant of transfected HEK293T cells with wild-type ADA2 or ADA2 variants T360A and N370K in homozygous conditions. Bar graphs represent percentage of ADA2 protein secretion relative to wild-type ADA2 100%. **C.** Quantification of ADA2 secretion of co-transfected HEK293T cells of ADA2 variants T360A and N370K together with wild-type ADA2 in heterozygous conditions. Bar graphs represent percentage of ADA2 secretion relative to wild-type ADA2 50%. **B-C.** Each bar represents mean ± SD from 3 independent experiments.

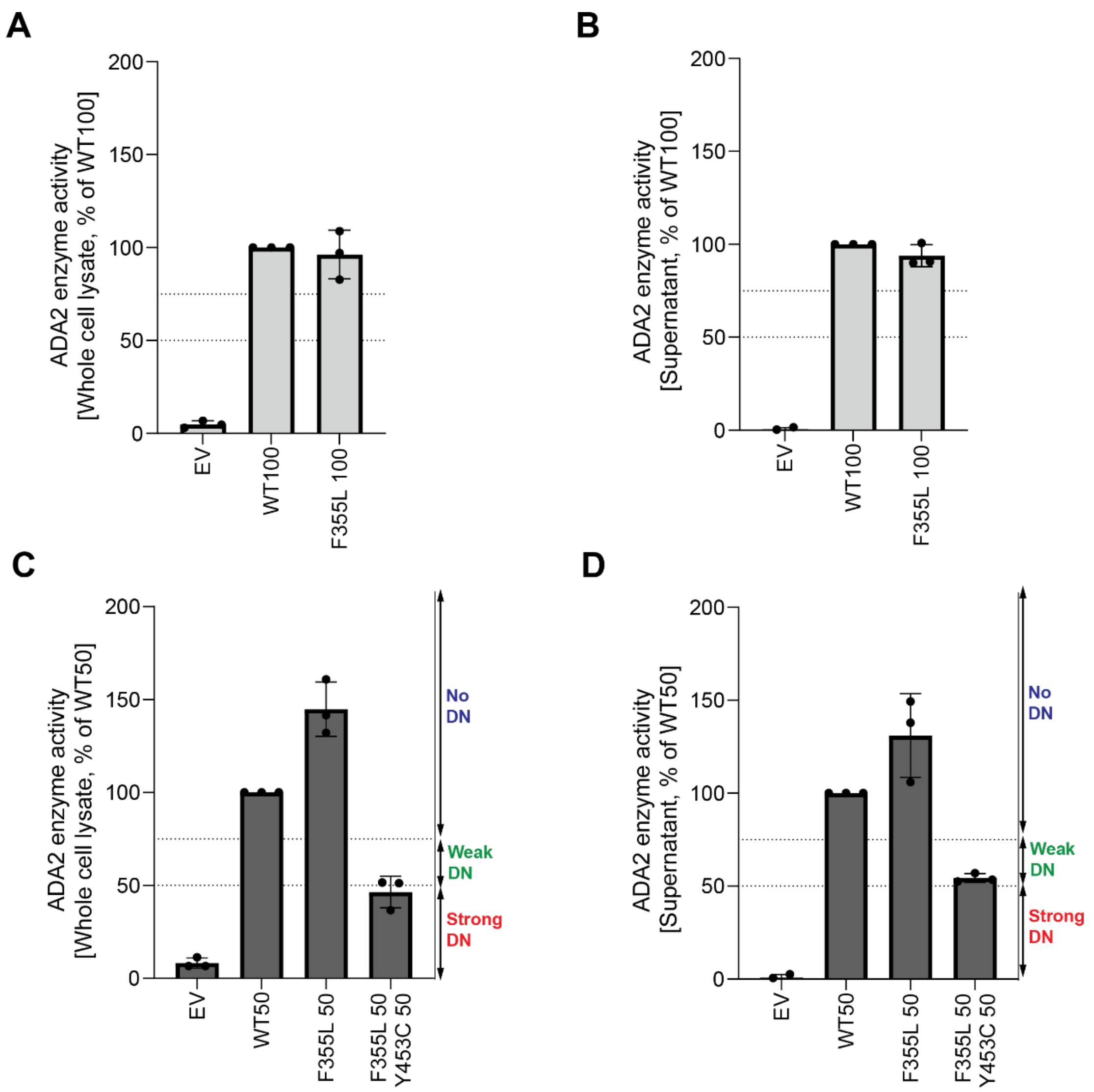


<u>Supplemental Figure 7. Adenosine deaminase activity of variant F355L in ADA2 in homozygous or heterozygous state.</u>

**A.** Adenosine deaminase activity in whole cell lysate of HEK293T transfected cells with WT and ADA2 variant F355L in homozygous conditions. Bar graphs represent the percentage of enzymatic activity relative to wild-type ADA2 100%. **B.** Adenosine deaminase activity in supernatant of HEK293T transfected cells with WT and ADA2 variant F355L in homozygous conditions. Bar graphs represent the percentage of enzymatic activity relative to wild-type ADA2 100%. **C.** Adenosine deaminase activity in whole cell lysate of HEK293T transfected cells with WT and/or ADA2 variant F355L in heterozygous conditions. Bar graphs represent the percentage of enzymatic activity relative to wild-type ADA2 50%.. **D.** Adenosine deaminase activity in supernatant of HEK293T transfected cells with WT and/or ADA2 variant F355L in heterozygous conditions. Bar graphs represent the percentage of enzymatic activity relative to wild-type ADA2 50%. **A-D.** Data represents mean ± SD from 3 independent experiments.

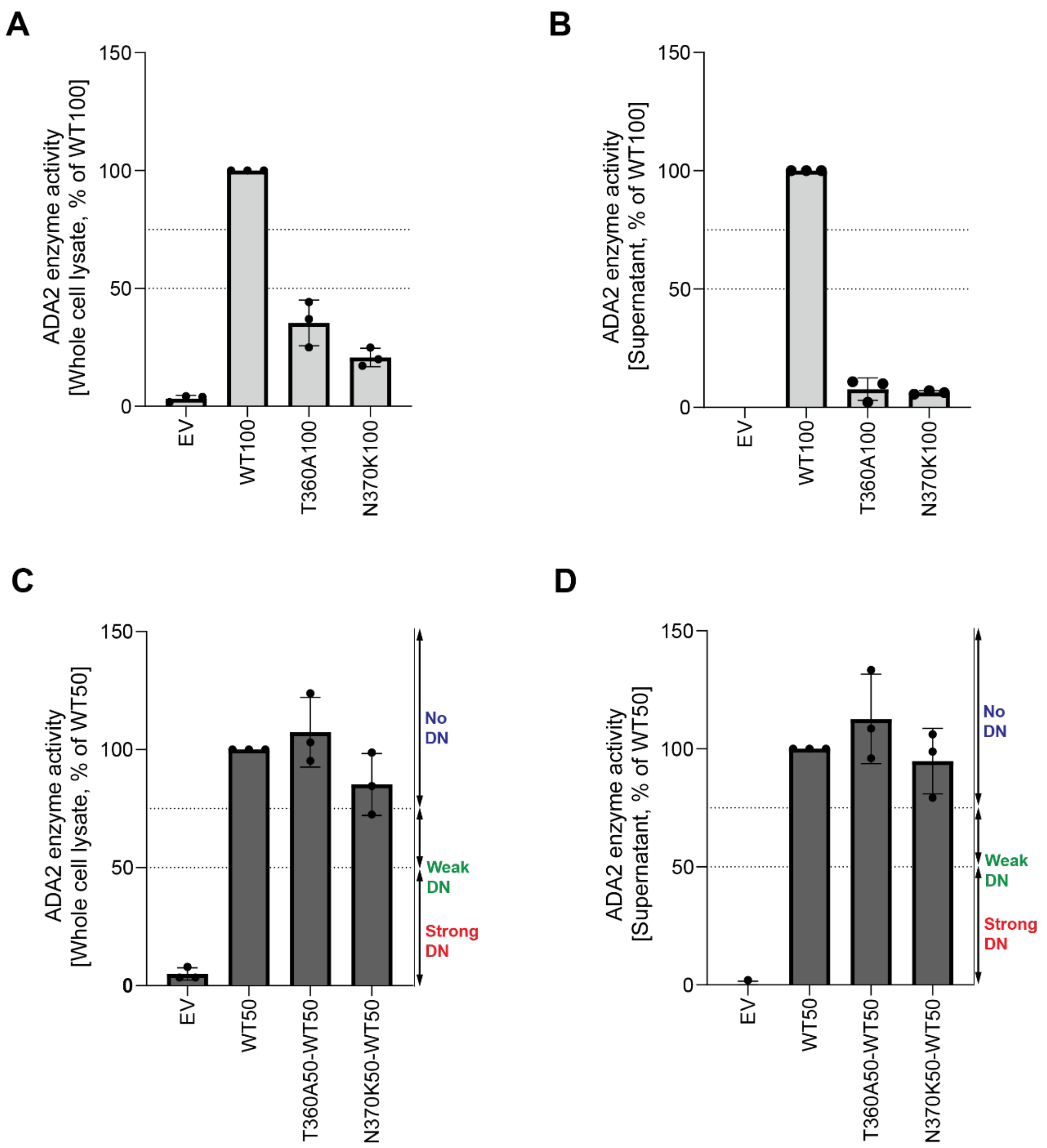


<u>Supplemental Figure 8. Adenosine deaminase activity of ADA2 variants T360A and N370K in homozygous or heterozygous state.</u>

**A.** Adenosine deaminase activity in whole cell lysate of HEK293T transfected cells with WT and ADA2 variants T360A and N370K in homozygous conditions. Bar graphs represent the percentage of enzymatic activity relative to wild-type ADA2 100%. **B.** Adenosine deaminase activity in supernatant of HEK293T transfected cells with WT and ADA2 variants T360A and N370K in homozygous conditions. Bar graphs represent the percentage of enzymatic activity relative to wild-type ADA2 100%. **C.** Adenosine deaminase activity whole cell lysate of HEK293T transfected cells with WT and/or ADA2 variants T360A and N370K in heterozygous conditions. Bar graphs represent the percentage of enzymatic activity relative to wild-type ADA2 50%. **D.** Adenosine deaminase activity in supernatant of HEK293T transfected cells with WT and/or ADA2 variants T360A and N370K in heterozygous conditions. Bar graphs represent the percentage of enzymatic activity relative to wild-type ADA2 50%. **A-D.** Data represents mean ± SD from 3 independent experiments.

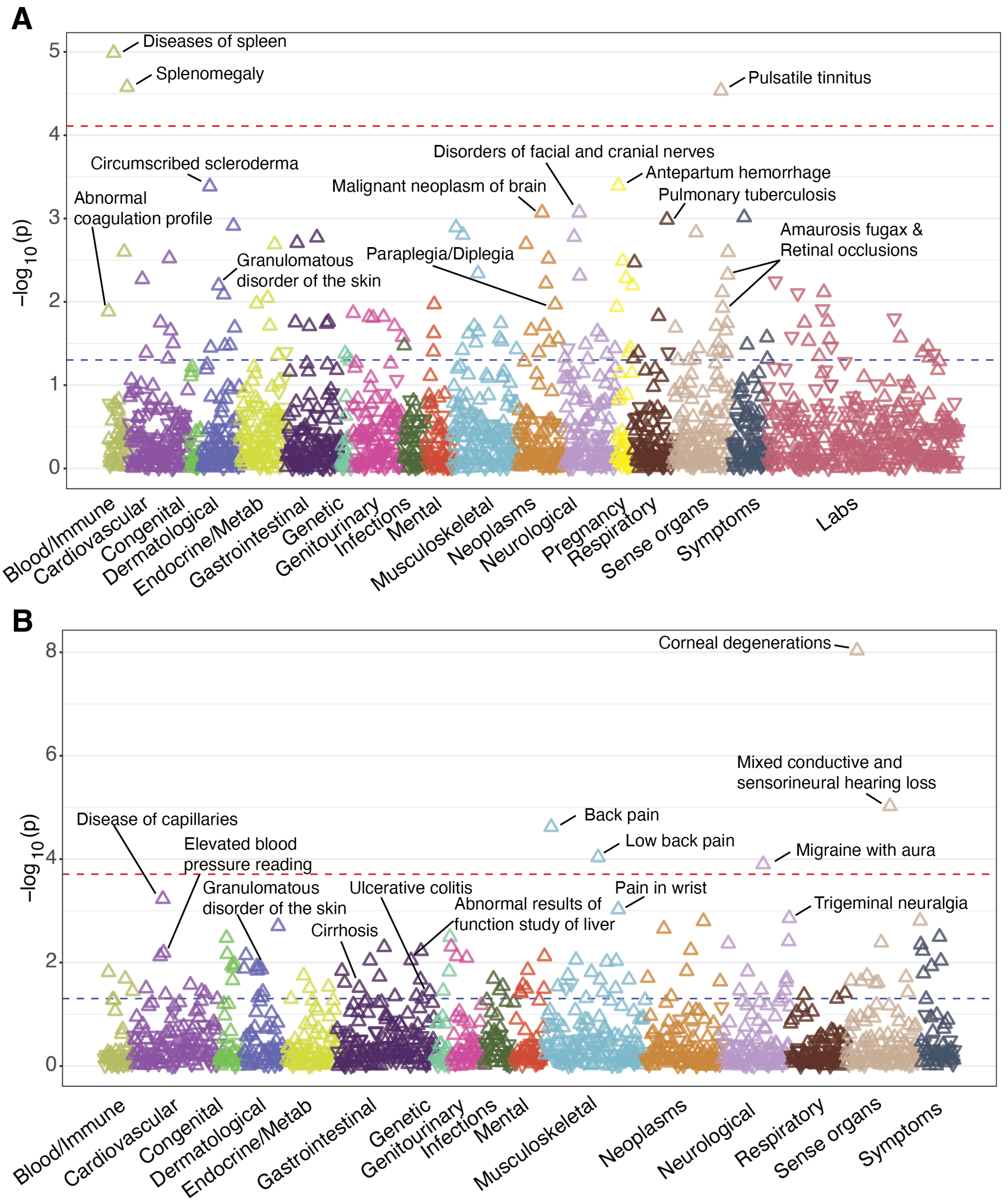


Supplemental Figure 9. PheWAS of *ADA2* pLOFs in the Bio*Me* BioBank and the UK Biobank.

**A.** Gene-based PheWAS results in the Bio*Me* BioBank (*n* = 27,742). **B.** Gene-based PheWAS results in the UK Biobank (*n* = 189,440). The direction of the triangles indicates the direction of effect (up: increased risk, down: decreased risk). The red dashed line represents false discovery rate (FDR)-adjusted *P* value threshold, whereas the blue dashed line indicates the nominal significance level (*P* = 0.05).
